# The genetic architecture of gene expression in individuals of African and European ancestry

**DOI:** 10.1101/2024.12.13.24318019

**Authors:** Kipper Fletez-Brant, Renan Sauteraud, Yanyu Liang, Steven Micheletti, Priyanka Nandakumar, Aarathi Sugathan, Kijoung Song, Taylor B. Cavazos, Amal Thomas, Robert J. Tunney, Barry Hicks, Jared O’Connell, Suyash Shringarpure, Katelyn Kukar, Meghan Moreno, Emily DelloRusso, Corinna D. Wong, Aaron Petrakovitz, Goutham Atla, Adrian Cortes, Padhraig Gormley, Laurence Howe, Rajashree Mishra, Daniel Seaton, the 23andMe Research Team, Robert C. Gentleman, Steven J. Pitts, Vladimir Vacic

**Author notes:** To whom correspondence should be addressed at, and. These authors contributed equally (Yanyu Liang, Steven Micheletti, Priyanka Nandakumar, Aarathi Sugathan).

## Abstract

We conducted two large scale studies of the genetics of gene expression in individuals of African ancestry within a cohort of consented 23andMe research participants and in LCL samples from the 1000 Genomes Project African superpopulation. We discovered nearly four times as many eQTLs, compared to tissue-matched eQTL studies in European cohorts. Additionally, we found that the majority of eQTLs were not detectable across populations; those that were, however, were found to be highly concordant. Performing eQTL studies in African ancestry cohorts resulted in more signals per gene and smaller credible sets of causal variants. We showed that comparisons of heritability of gene expression could be confounded by population substructure, but that variation in local genetic ancestry did not majorly impact eQTL discovery. Finally, we showed improvements in variant-to-gene mapping of African-American GWAS signals when using African compared to European ancestry eQTL studies

## Introduction

Underrepresentation of individuals of African ancestry in biomedical research is a serious ethical issue leading to healthcare disparities worldwide, and a major missed scientific opportunity to understand the genetic basis of disease . Initiatives like H3Africa^2^, Southern African Human Genome Programme^3^ and MalariaGen^4^ have conducted genomic studies with research participants from African countries, while the All of US^5^ and Million Veteran Program (MVP)^6^ have performed large GWAS in African-American cohorts in the United States. While associations can be identified more easily in African compared to non-African populations as a result of increased genetic diversity^7^, interpreting GWAS results and mapping disease risk genes requires a functional link between genetics and an intermediate molecular phenotype, such as expression quantitative trait loci (eQTL). Although eQTL consortia such as GTEx^8^ ncluded African-American samples and performed follow-ups that focused on analysis of these samples^9^, eQTL studies specifically conducted in African^10,11^ or African-American cohorts^12,13^ as well as QTL studies broadly^14^, are fewer. Comparative studies of regulatory variation in diverse human populations in the HapMap3 cohort demonstrated significant sharing of *cis-*regulatory variation across populations, and for shared eQTLs, near-perfect concordance of directionalities and effect sizes^15^. However, differences in allele frequencies across populations cause differential eQTL discovery power^15^ and limit transferability of gene expression prediction models^16^. Genetic European Variation in Disease (GEUVADIS)^10^, the first large RNA-seq based cross-population study of regulatory variation, highlighted differences in transcript usage and differences in overall gene expression, but concluded that these two were largely mediated by separate genetic variants. eQTL analyses in the Human Genome Diversity Panel^17^ cohort reported that 25% of variation in expression was attributable to ancestry, and that 76% of this variation is due to expression rather than splicing. A recent study of eQTLs in African-American and Latino populations linked heritability of gene expression and population heterozygosity, and showed prevalence of population specific eQTLs to be 30% and 8% within ancestral African and indigenous American genomic segments respectively^13^. African Functional Genomics Resource (AFGR) compiled gene expression measured in 1000 Genomes Project African samples and additional Maasai individuals, as well as open chromatin in a subset of 100 individuals, and assembled a comprehensive dataset of expression, splicing and chromatin accessibility QTLs^18^

To address underrepresentation of individuals of African ancestry (AFR) in research, we conducted eQTL studies in two different cohorts. We recruited a cohort of consented 23andMe research participants, which we refer to as the Black Representation in Genomic Research (BRGR) study. We collected saliva and venous blood samples for whole-genome sequencing (WGS) and RNA-seq, respectively, from 737 individuals. Additionally, we sequenced RNA extracted from 659 lymphoblastoid cell lines (LCLs) belonging to the African ancestry superpopulation in the 1000 Genomes Project^19^, comprised of individuals from continental Africa (including GEUVADIS Yoruba individuals^10^ and most AFGR samples^18^) and admixed individuals from the African diaspora. Using publicly available WGS of these individuals^20^, we analyzed the genetics of gene expression in LCLs (Fig. 1a). As comparisons, we analyzed eQTLs in venous blood and LCLs from two large European cohorts^10,21,22^ (Table 1) and subsequently investigated sharing of eQTLs. We used AFR and EUR eQTLs to annotate African-American GWAS signals in 6 studies from MVP^23–25^, 11 studies from the Blood Cell Consortium (BCX)^14^ and in the 23andMe GWAS of height in African-Americans.

**Figure 1.**
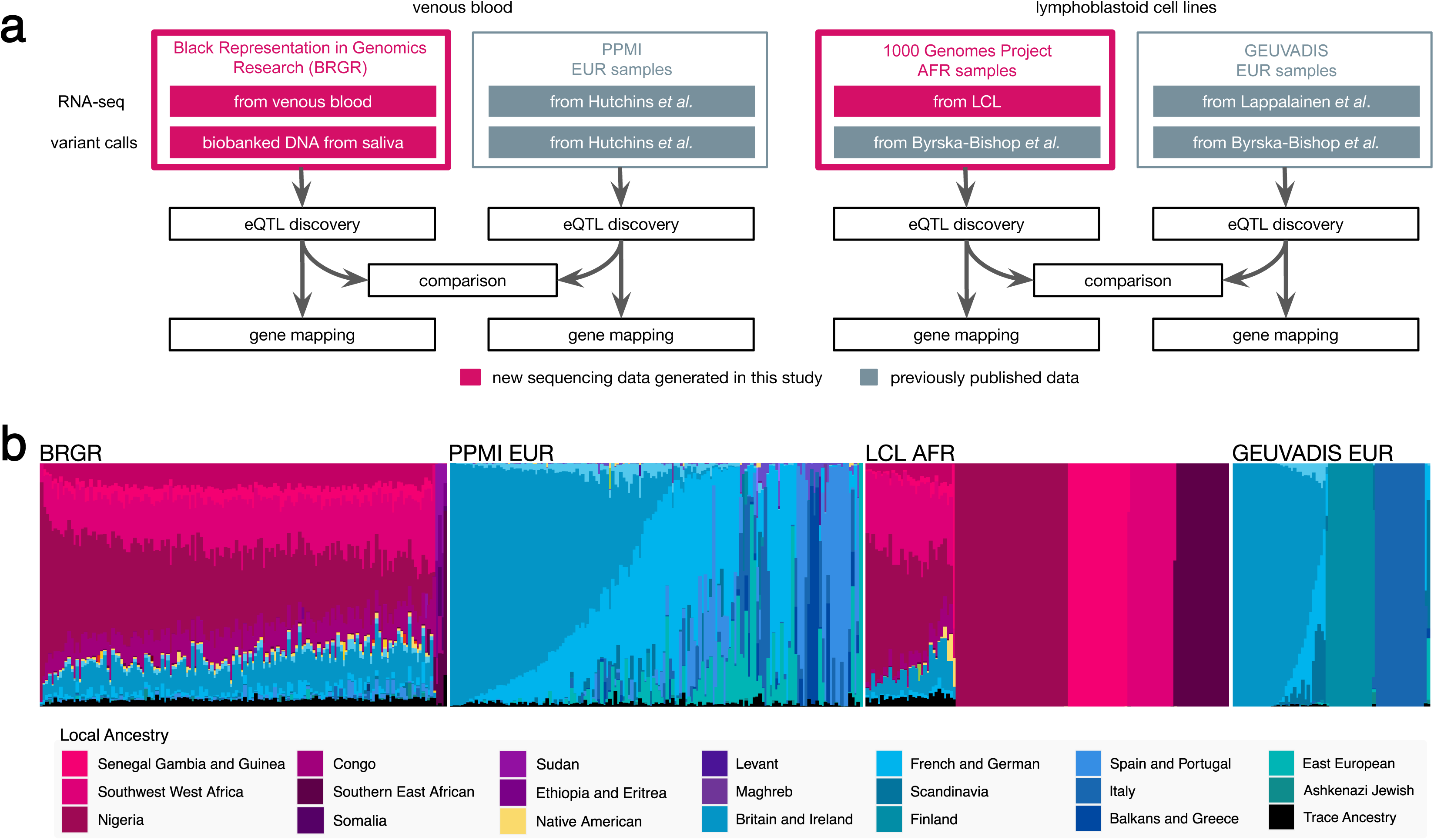
Schematic diagram of the study design and Ancestry Composition estimates for each eQTL cohort. **(a)** Schematic diagram of the four eQTL study arms. Fuchsia-colored boxes indicate new RNA-seq or WGS data generated for this study. **(b)** Results of the Ancestry Composition algorithm applied to the four eQTL cohorts. Each vertical line represents genome-wide proportions for a single individual.

**Table 1.** Summary of the four cohorts and eQTLs discoveries. eQTLs are defined as (eVariant, eGene) pairs, eVariants as variants associated with a change in gene expression of one or more protein-coding eGenes, and eGenes as protein-coding genes having one or more eQTLs.

| Cell line / tissue | Venous blood |  | LCL |  |
| --- | --- | --- | --- | --- |
| Dataset | BRGR | PPMI | LCL_AFR | GEUVADIS |
| Population | AFR | EUR | AFR | EUR |
| Sample size | 737 | 752 | 659 | 358 |
| Total variants in cohort | 15,598,612 | 8,672,989 | 15,585,478 | 8,895,186 |
| Genes tested | 14,162 | 17,233 | 13,584 | 14,624 |
| eGenes discovered | 10,044 (70.9%) | 4,277 (24.8%) | 8,843 (65.1%) | 2,527 (17.3%) |
| eQTLs discovered | 21,130 | 5,661 | 16,265 | 2,904 |
| eVariants discovered | 20,295 | 5,380 | 15,832 | 2,843 |

## Results

### eQTL cohorts capture African and European ancestral backgrounds

We applied 23andMe’s Ancestry Composition (AC)^26^ algorithm to genetic variants called from WGS data in the four cohorts and confirmed African and European ancestry components (Fig. 1b). Plurality of BRGR participants are from the South Census Region (Suppl. Fig. 1, Suppl. Table 1). Genome-wide AC proportions of BRGR participants are similar to previously published ancestry of individuals of African descent in the U.S.^27^, with high representation of Sub-Saharan African ancestry (μ^African^=0.80±0.11, mean±sd) and moderate representation of European ancestry (μ^European^=0.17±0.10, Suppl. Table 2). Plurality of local African ancestry is Nigerian (μ^Nigerian^=0.29±0.09), with the majority of individuals grouping closely to the Igbo reference group according to a graph method based on shared identical-by-descent (IBD) segments^28^ (Suppl. Fig. 2a). Additionally, the distribution of ancestry found in the BRGR cohort is representative of all genotyped African-American customers in the 23andMe database (based on a randomization test, Suppl. Table 2). The Parkinson Progression Marker Initiative (PPMI)^22,29^ dataset was predominantly European (μ^European^=0.97±0.09), with the plurality of local European ancestry being British and Irish (μ^British-Irish^=0.35±0.37), followed by Iberian (μ^Iberian^=0.13±0.31). For the 1000 Genomes Project eQTL cohorts, our genome-wide AC proportions recapitulate established population genetics results^19^ (Fig. 1b, Suppl. Table 2,3).

### Increased genetic diversity improves eQTL discovery power

RNA-seq from each of the four datasets underwent the same QC and gene expression quantification procedures, and yielded comparable numbers of protein-coding genes for eQTL testing (slightly more in PPMI, see Table 1). Approximately 15.6M variants pass QC filters in each African cohort compared to 8.6M variants in European cohorts, in accordance with the known larger number of variants per genome in individuals of African ancestry^19^. Comparing MAF spectra across ancestry groups reveals enrichment of rare variants in African versus tissue-matched European cohorts (Fig. 2a, “all variants”).

**Figure 2.**
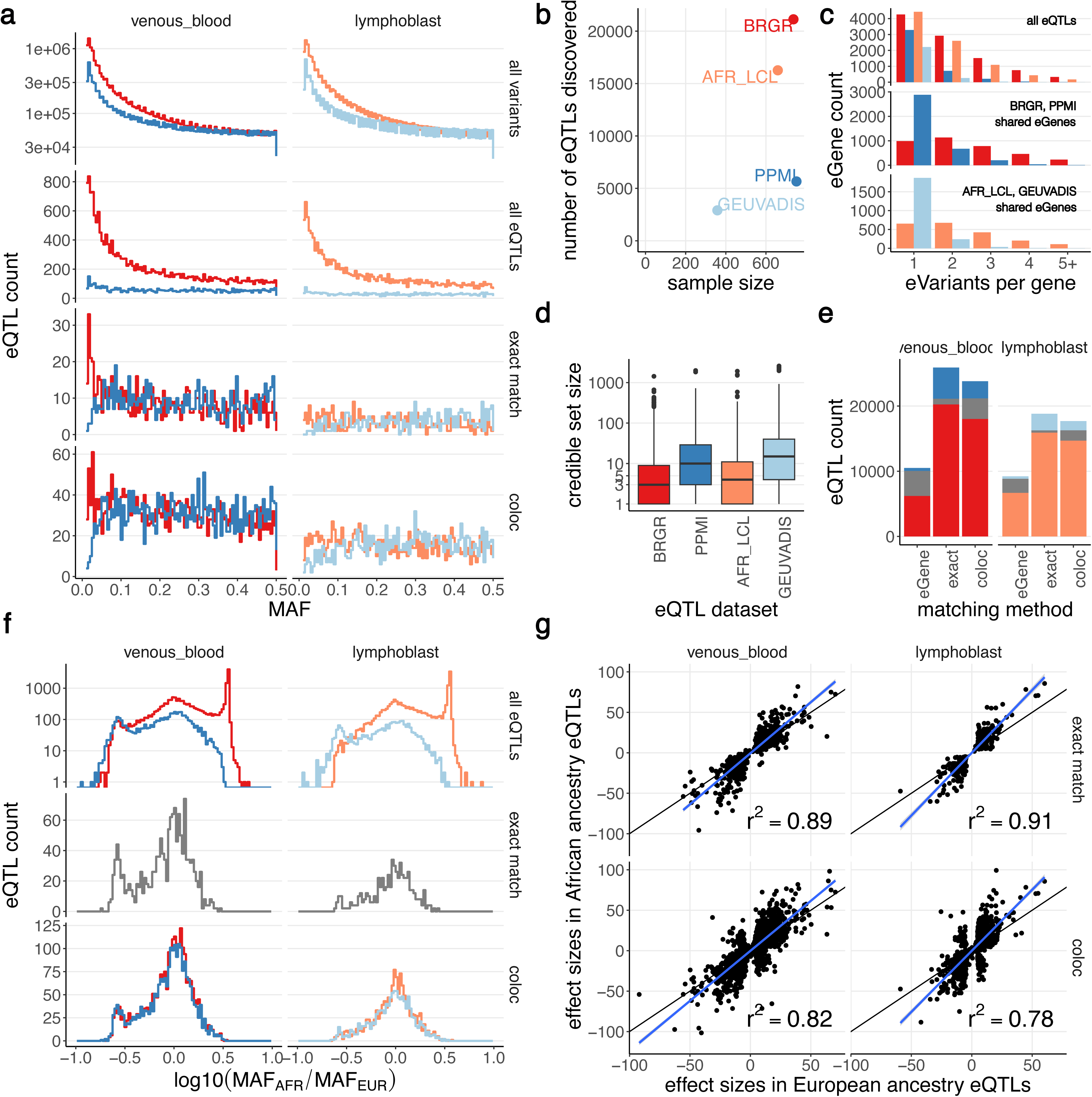
Statistics of eQTL calls and eQTL sharing across cohorts. **(a)** Minor allele frequency histograms of all variants and all detected eQTLs, as well as for eQTLs that were also observed in the matching tissue in the complementary population. **(b)** Scatter plot of number of eQTLs discovered against cohort size shows higher numbers of eQTLs in African ancestry groups even when normalized by study size. **(c)** Distributions of numbers of distinct eVariants per eGene show a higher number of eVariants in AFR cohorts. **(d)** Distributions of credible set sizes have smaller medians in European ancestry datasets. **(e)** Sharing of eGenes and eQTLs between African and European studies in the same tissue or cell type is broadly low, irrespective of the method of comparison. **(f)** Spectra of log10(MAF_AFR_ / MAF_EUR_) for eQTLs from African (red) and European (blue) ancestry cohorts, in venous blood and LCLs respectively. **(g)** Shared eQTLs are largely concordant in their direction of effect (Pearson r^2^ 0.89-0.91 for exactly the same eVariants, r^2^ = 0.79-0.83 for colocalizing eVariants), with on average slightly elevated magnitude of effect in African cohorts (blue line is best linear fit, black line x=y for comparison).

To discover eQTLs, we applied SuSiE^30,31^ to individual level data from each study, using default parameters. For each discovered eGene SuSiE returns independent eQTL signals as a collection of credible sets (CSs). Within each CS, we reported the variant with the largest posterior inclusion probability (PIP) as the index eVariant. In BRGR (sample size n=737) we detected 21,130 eQTLs regulating 10,044 eGenes via 20,295 eVariants, compared to 5,661 eQTLs (4,277 eGenes; 5,380 eVariants) in PPMI (n=752; Table 1; Fig. 2b). In LCL_AFR (n=659) we found 16,265 eQTLs (13,584 eGenes; 15,832 eVariants) compared to 2,904 eQTLs (2,527 eGenes; 2,843 eVariants) in GEUVADIS (n=358), however we here note the differential sample size in LCLs. MAF spectra (Fig. 2a, “all eQTLs”) showed that eQTLs called in African cohorts are enriched for rare variants, compared to a lack in European eQTLs. MAF relative increase in AFR compared to EUR is bimodal in both cell types, with a majority of eQTLs (11,662 in BRGR and 10,032 in LCL_AFR) with at least 3.16x (log_10_ ratio = 0.5) greater MAF (Fig. 2f, “all eQTLs”).

We discovered fewer eVariants per eGene in Europeans, with 77% of PPMI and 88% of GEUVADIS eGenes having only one eVariant. Both African cohorts have comparable trends in numbers of eVariants per eGene (Fig. 2c) and CS sizes are two times smaller in African compared to European cohorts (Fig. 2d).

### Majority of eQTLs are population- and tissue-specific, but the effects of shared eQTLs are concordant

We first assessed sharing of eQTLs conservatively defined as exactly matching eGene and eVariant. We found 872 eQTLs shared between BRGR and PPMI, and 341 between LCL_AFR and GEUVADIS. As fraction of constituent datasets, eQTL sharing was infrequent (15.4% of PPMI, 4.1% of BRGR eQTLs; 2.1% of LCL_AFR, 11.7% of GEUVADIS eQTLs; Fig. 2e). However, shared eQTLs exhibited consistent effect sizes (Fig. 2g), with Pearson correlation r^2^ 0.89 (p-value<2.2×10^-16^) and 0.91 (p-value<2.2×10^-16^) for venous blood and LCL datasets, respectively. Shared eQTLs had smaller CSs (Suppl. Fig. 3), and 75% of CSs in African cohorts contained 1 or 2 variants, while in PPMI 75% of CSs had up to 7 variants, and over 10 in GEUVADIS. Shared eQTLs had similar MAF spectra, except for rare alleles, which were underrepresented in Europeans (Fig. 2f). Shared venous blood eQTLs either had comparable or greater MAF in AFR, while LCL eQTL MAFs were comparable. Within-population cross-tissue sharing was rare: 1,987 eQTLs were shared within African datasets (9% BRGR; 12% LCL_AFR), while 176 eQTLs were shared within European datasets (3% PPMI; 6% GEUVADIS).

Next, we considered eQTLs shared if eGenes matched and pairs of signals colocalized^32^ with posterior probability H_4_ ≥ 0.5. We found that 55.5% of eQTLs identified in PPMI were shared with BRGR, and 14.9% conversely (Table 2). 54% of eQTLs identified in GEUVADIS were shared with LCL_AFR and 9.6% conversely. eQTL effect sizes were consistent, with r^2^=0.83 (p-value<2.2×10^-16^) and 0.79 (p-value<2.2×10^-16^) for venous blood and LCLs, respectively. Across tissues, African cohorts shared 4,545 eQTLs (22% of BRGR and 27% of LCL_AFR eQTLs, respectively) while European cohorts shared 769 eQTLs (14% of PPMI and 26% of GEUVADIS eQTLs, respectively). CSs of colocalized eQTLs were smaller in African than in European cohorts (Suppl. Fig. 3) and shared eQTLs have comparable MAF. Venous blood specifically shows a striking enrichment in BRGR eQTLs of eVariants more common in BRGR versus PPMI (Fig. 2f, x-axis=+0.5), and also that variants more common in PPMI (compared to BRGR) are detected in BRGR (Fig. 2f, x-axis=-0.5). The converse is not true: variants more prevalent in BRGR are not detected in PPMI, although PPMI is also enriched for variants more common in PPMI.

**Table 2.** Summary of eQTLs and eGenes overlap across populations and tissues. Replicating here means that (eGene, eVariant) pair from the first dataset appeared in one of the credible sets for the same eGene in the second dataset. We report both directions of replication (1^st^ replicating in 2^nd^; 2^nd^ replicating in 1^st^) separately. Where applicable, numbers in square brackets are values of Jaccard similarity coefficient defined as the size of the intersection divided by the size of the union. For replication, numbers in square brackets are % of hits replicating. Numbers and % in parentheses indicate eQTLs with a common directionality of effect. For exact eVariant match and replication, direction was based on the sign of coefficient of the eVariant; for coloc, direction was based on the sign of the z-score correlation coefficient.

|  | BRGR and<br>PPMI<br>(both venous<br>blood) | LCL_AFR and<br>GEUVADIS<br>(both LCL) | BRGR and<br>LCL_AFR<br>(both AFR) | PPMI and<br>GEUVADIS<br>(both EUR) |
| --- | --- | --- | --- | --- |
| Exact eVariant<br>match | 872 [0.03]<br>(866; 99%) | 341 [0.02]<br>(341; 100%) | 1,987 [0.06]<br>(1,938; 98%) | 176 [0.02]<br>(168; 95%) |
| Colocalizing<br>eQTLs ($H_4 \geq 0.8$) | 2,772 [0.11]<br>(2,534; 91%) | 1,330 [0.07]<br>(1,199; 90%) | 4,161 [0.10]<br>(3,858; 93%) | 656 [0.08]<br>(558; 85%) |
| Colocalizing<br>eQTLs ($H_4 \geq 0.5$) | 3,145 [0.13]<br>(2,820; 90%) | 1,569 [0.09]<br>(1,398; 89%) | 4,545 [0.14]<br>(4,159; 92%) | 769 [0.10]<br>(630; 82%) |
| 1<sup>st</sup> replicating in<br>2<sup>nd</sup> | 4,508 [21%]<br>(4,408; 98%) | 2,621 [16%]<br>(2,598; 99%) | 3,406 [16%]<br>(3,279; 96%) | 836 [15%]<br>(763; 91%) |
| 2<sup>nd</sup> replicating in<br>1<sup>st</sup> | 1,295 [23%]<br>(1,283; 99%) | 511 [18%]<br>(511; 100%) | 3,405 [21%]<br>(3,278; 96%) | 536 [18%]<br>(492; 92%) |
| Shared eGenes | 3,822 [0.37] | 2,153 [0.23] | 6,843 [0.57] | 1,331 [0.24] |

When comparing MAF spectra of shared (either exact match or coloc) and unshared eQTLs for a given tissue, all shared eQTLs have either comparable or larger MAF in EUR versus AFR (Fig. 2f). eQTLs with meaningfully larger allele frequencies in individuals of African ancestry tend to be unshared, highlighting the value of conducting eQTL studies in diverse cohorts.

We next measured replication, which within the SuSiE framework we defined as matching eGenes and the variant with the largest PIP in one CS (aka eVariant) observed in the other CS (Table 2). 23% of eQTLs found in PPMI replicated in BRGR and 21% vice versa, while for LCL cohorts, 18% of GEUVADIS eQTLs replicated in LCL_AFR and 16% vice versa. Comparing tissues, 16% of BRGR eQTLs were found in LCL_AFR and 21% vice versa, and 15% of PPMI eQTLs were found in GEUVADIS and 18% vice versa.

Finally, we compared detected eGenes, irrespective of eVariants or signal colocalization. 3,822 eGenes were shared between BRGR and PPMI (respectively, 27% and 22% of genes). 2,153 eGenes were shared between LCL_AFR and GEUVADIS (5.8% and 14.7% of genes). 6,843 of eGenes were shared between BRGR and LCL_AFR (48% and 50% of eGenes, respectively) while 1,331 eGenes were shared PPMI and GEUVADIS (7% and 9% of eGenes, respectively). In summary, our results indicated that only a small fraction of discovered eQTLs were shared across populations.

### Heritability analyses of gene expression are confounded in cross-ancestry comparisons

We measured *h^2^* of gene expression in BRGR and PPMI using GCTA-GREML^33^ and identified 9,777 eGenes that showed significant heritability in both cohorts. Average heritability in BRGR is significantly higher (*h^2^*=0.31) than in PPMI (*h^2^*=0.20). Out of the 9,777, we identified a subset of 1,016 eGenes with eQTL signals that colocalize across cohorts and measured the relationship of differential genetic variance of index SNPs to differential *h^2^* (Δ*h^2^*) across genes. Despite the assumption of shared causal variants for signals that colocalize across cohorts, we find that Δ*h^2^*, the difference in heritability, is positively associated with differential genetic variance (*p-value*=2.9×10^-43^), accounting for 17% of variation in Δ*h^2^* (Fig. 3b, Suppl. Fig. 4a). We also find that the ratio of unexplained to heritable variation, defined as *q = h^2^* / (1-*h^2^)* is associated with differential genetic variance of eVariants (Fig. 3c; Suppl. Fig. 4b,c; Suppl. Table 4), indicating that comparisons of heritability of shared casual signals are confounded by differential MAF.

**Figure 3.**
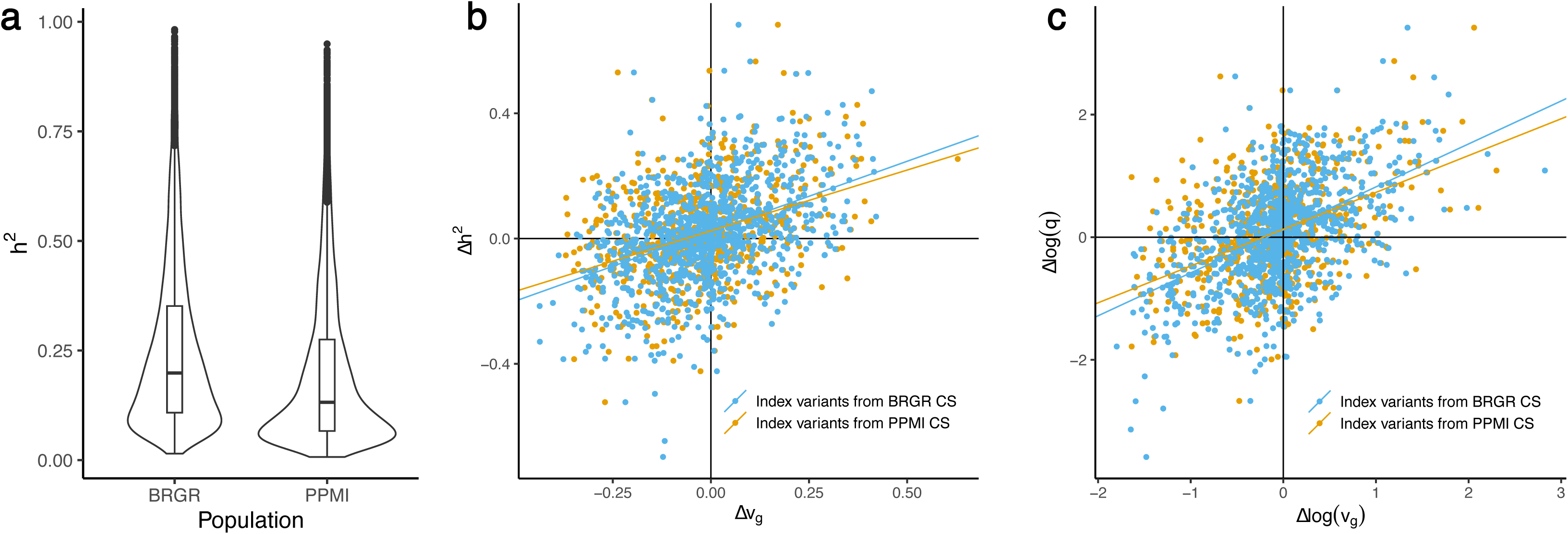
Cis-heritability of gene expression in venous blood and difference in genetic diversity explaining population differences. **(a)** Comparison of h^2^ across populations (BRGR in blue and PPMI in yellow) showing a significant difference in h^2^ (p-value < 2.2 10^-16^). **(b), (c)** Among the 183 genes whose credible sets are colocalized between BRGR and PPMI, we define the causal eQTLs as the index variants from BRGR (blue) or PPMI (yellow) credible sets and calculate the genetic diversity v_g_ of the causal eQTLs in BRGR and PPMI respectively. To examine the relation between genetic diversity and heritability, we introduce a transformed heritability, q = h^2^ / (1 - h^2^). **(b)** For each of the 183 genes, differences in genetic diversity ΔV_g_ = V_g,BRGR_ - V_g,PPMI_ are shown on x-axis and differences in heritability 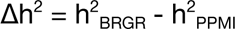 shown on y-axis. The line is drawn from the linear fit Δh^2^ ∼ 1 + Δv_g_ **(c)** For each of the 183 genes, differences in logarithm of genetic diversity between BRGR and PPMI (Δlog(v_g_) = log(v_g, BRGR_) - log(v_g, PPMI_)) are shown on x-axis and differences in logarithm of transformed heritability (Δlog(q) = log(q_BRGR_) - log(q_PPMI_)) are shown on y-axis. The line is drawn from the linear fit Δlog(q) ∼ 1 + Δlog(v_g_).

### Standard best practices in eQTL calling adequately control for ancestry as confounder

To assess unmodeled effects of ancestry on our findings in eQTL sharing and heritability, we compared the contribution of global ancestry using the standard approach of genetic PC covariates to modeling local ancestry using Tractor^34^ and found that 80% of eGenes were identified in both models (Suppl. Fig. 5a). Another 15% of eGenes were significant in both models but with different eVariants. As Hou *et al* ^35^ we found that Tractor was well-powered to identify effect size heterogeneity by ancestry and underpowered otherwise (Suppl. Fig. 6b), and for the majority of eQTLs we did not observe effect heterogeneity. We concluded that individual eQTL datasets were not impacted by ancestry-induced biases.

### Genetic determinants of Duffy-null associated neutrophil count connect gene expression to cellular and physiological phenotypes

Reasoning that blood cell phenotypes with greater prevalence in Africans compared to Europeans should have detectable correlates in BRGR eQTLs, we investigated eQTL signals for Duffy-null associated neutrophil count (DnANC), a phenotype characterized by lower neutrophil and leukocyte counts without increased infection risk^36,37^ DnANC is observed in individuals of African, Middle Eastern, and West Indian descent, with estimated prevalence as high as 25-50% in AFR^38^. DnANC is driven by the Duffy-null genotype CC of rs2814778 (ClinVar:18395) n *ACKR1*^37,39^. Individuals with Duffy-null allele have increased protection against *Plasmodium vivax* malaria infection and increased susceptibility to HIV-1 trans-infection^40^ *ACKR1* venous blood expression correlates with neutrophil counts because of its effects on neutrophil homeostasis; however *ACKR1* is a chemokine scavenger receptor expressed in erythrocytes and endothelial cells, not neutrophils.

We found multiple eQTLs for *ACKR1* in the BRGR and PPMI cohorts (Fig. 4): rs2814778 is an eQTL in the BRGR study (*β*±se*=-O.96±O.O2,* p-value<2.2×10^-308^), is more common in Africans (MAF_AFR_=0.16, MAF_EUR_=0.002, based on gnomAD^41^ v4) and is located in the promoter of *ACKR1*. In BCX, rs2814778 has the strongest association of any blood-cell trait in African-ancestry individuals, and is associated with counts of neutrophils (β=0.86±0.02, p-value=3.99×10^-432^), white blood cells (β=0.70±0.02, p-value=1.02×10^-330^) and monocytes (β=0.32±0.02, p-value=1.92×10^-63^). We also identified rs863005 as a secondary *ACKR1* eQTL in the BRGR cohort. We detected rs12075 as an eQTL for *ACKR1* in PPMI. This variant is more common in Europeans (MAF_AFR_=0.07, MAF_EUR_=0.42) and is a QTL for monocyte (β=0.027±0.002, p-value=3.24×10^-41^) and basophil counts (β=0.028±0.002, p-value=6.63×10^-39^) in Europeans in BCX. rs12075 is a coding variant for *ACKR1*^42^ and has been reported as a regulator of the monocyte chemokine *MCP-1*^43^. This European eQTL colocalizes (H_4_=0.6) with the rs863005 eQTL from BRGR. We didn’t detect any eQTLs for *ACKR1* in the LCL datasets, as this gene is not expressed in B cells^44,45^

**Figure 4.**
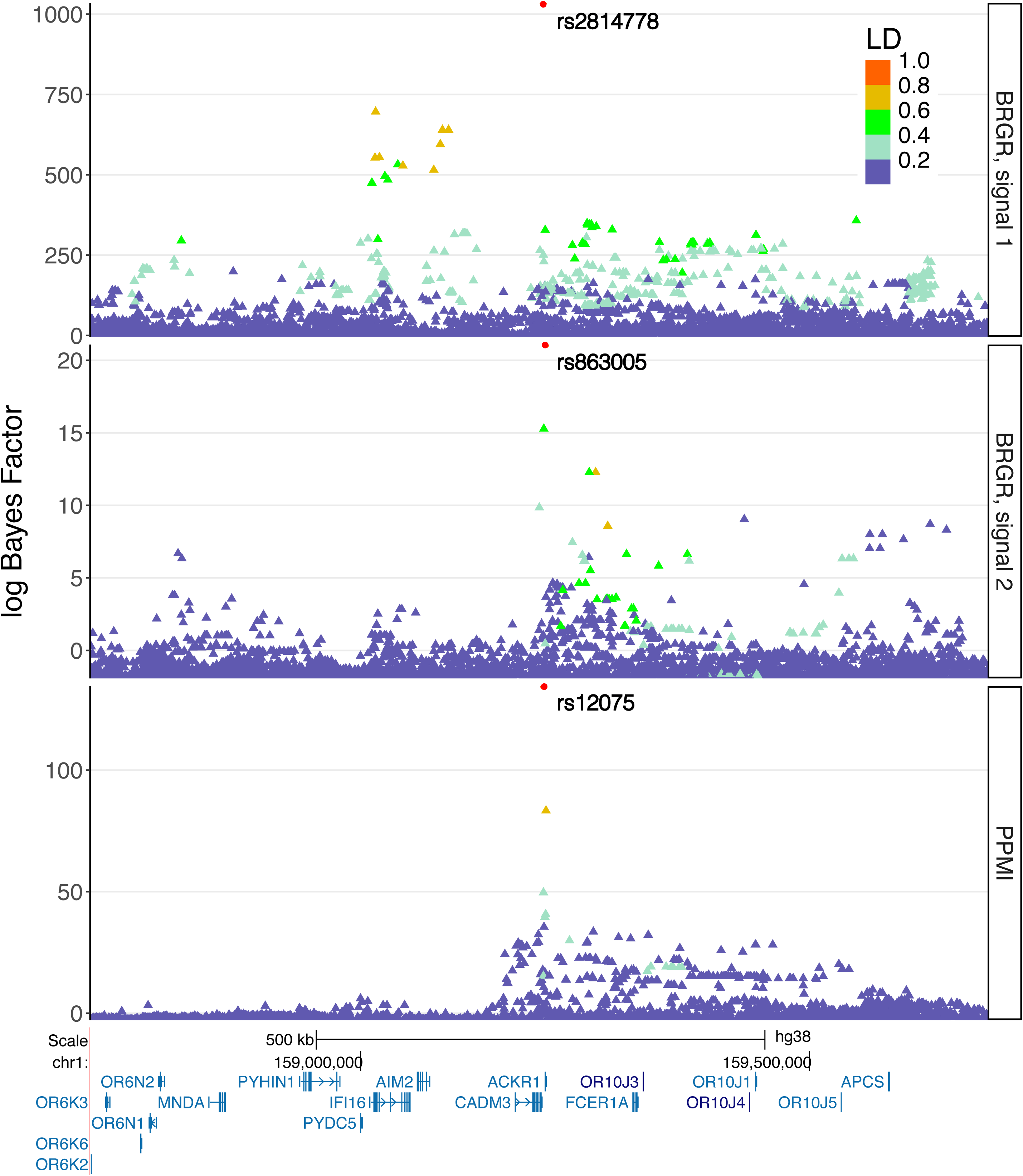
*ACKR1* eQTLs in BRGR and PPMI, colored by in-sample LD to highest logBF variant. Shown are SuSiE log Bayes factors (logBFs) for each SNP in a given eQTL signal. logBFs reflect the evidence that a SNP is the causal SNP, and are proportional to posterior inclusion probabilities. BGRG signal #1 (PIP = 1.0), with the Duffy null allele variant rs2814778 as the eVariant. BRGR signal #2 (PIP = 0.996), with rs863005 as the eVariant. Compared to signal #2, #1 exhibits a longer segment of variants in linkage disequilibrium with each other, which may reflect the selective pressure specifically on the haplotype carrying the CC allele of rs2814778. PPMI signal for rs12075 (PIP = 0.999). The signal is localized to the eVariant, and is in linkage disequilibrium with few nearby variants.

### African ancestry eQTLs improve annotation of African-American GWAS

To assess the utility of African ancestry eQTL datasets for annotating African-American GWAS, we mapped associations in 22 GWAS performed in African-American cohorts: 8 cardio-metabolic traits from the Million Veteran Program (MVP), 13 blood measurement traits from the Blood Cell Consortium (BCX) and 23andMe’s GWAS of height (Suppl. Table 5). Across the board, using AFR eQTLs lead to higher fractions of African-American GWAS signals annotated with at least one gene mapping hypothesis. In MVP, BCX and height studies BRGR eQTLs mapped 1.3-3x more signals to genes compared to PPMI. In BCX and height studies, LCL_AFR eQTLs mapped 1.75-2.77x more signals than GEUVADIS. GEUVADIS eQTLs didn’t contribute to gene mapping in MVP while LCL_AFR eQTLs mapped 1.3% of association signals to genes (Table 3). BRGR eQTLs map at least one GWAS hit per phenotype in 71.4% of BCX phenotypes and 80% of MVP phenotypes, compared to 42.9% and 40% for PPMI respectively. Similarly, LCL_AFR eQTLs annotate at least one GWAS hit per phenotype in 50% of BCX and 60% of MVP phenotypes compared 35.7% and 0% for GEUVADIS. All four eQTL datasets annotated at least one GWAS hit in 23andMe height.

**Table 3.** Summary of variant-to-gene mapping of African-American GWAS signals. MVP - the VA Million Veteran Program; BCX - Blood Cell Consortium. Detailed per-phenotype GWAS breakdown is provided in Suppl. Table 5.

| GWAS dataset | GWAS signals | H<sub>4</sub> cutoff | Number of GWAS-to-eQTL signal colocalizations |  |  |  | r<sup>2</sup> with coding |
| --- | --- | --- | --- | --- | --- | --- | --- |
|  |  |  | BRGR AFR | PPMI EUR | LCL_AFR AFR | GEUVADIS EUR |  |
| MVP | 312 | 0.8 | 6 | 2 | 4 | 0 | 15 |
|  |  | 0.5 | 9 | 2 | 4 | 1 | 16 |
| BCX | 461 | 0.8 | 32 | 12 | 14 | 8 | 5 |
|  |  | 0.5 | 41 | 16 | 19 | 9 | 5 |
| Height | 810 | 0.8 | 32 | 22 | 26 | 8 | 21 |
|  |  | 0.5 | 52 | 35 | 46 | 17 | 37 |

eQTLs discovered in BRGR and not shared with PPMI contributed 50 gene-phenotype hypotheses relating 38 GWAS signals in 15 phenotypes to 40 genes, while only 9 geno-phenotype pairs (9 GWAS signals in 3 phenotypes and 9 genes) were discovered with eQTLs unique to PPMI. eQTLs discovered in LCL_AFR and not shared with GEUVADIS contributed 34 gene-phenotype hypotheses (31 GWAS signals in 8 phenotypes, 32 genes), while only 4 were discovered with eQTLs unique to GEUVADIS (4 GWAS signals in 2 phenotypes, 4 genes).

We next explored the novelty of our gene-phenotype hypotheses in OpenTargets Genetics v22.10, regardless of the specific eQTL or discovery tissue or cohort. Of the gene-phenotype hypotheses contributed by eQTLs only discovered in BRGR, 20 out of 50 hypotheses have not been previously reported. Notably this includes thalassemia variant rs33930165 (Clinvar ID: 15126) and the hemoglobin gene *HBD* and BCX phenotypes MCHC, RBC, RDW (all H_4_=1.0; see Suppl. Table 6). Of those that have a close matching phenotype in OpenTargets — e.g. *TRIP10* and BCX phenotype mean platelet volume (MVP)^46^; max H_4,OpenTargets_=0.37 and H_4, BRGR_=0.99 — the average relative increase of H_4_ under African eQTLs was 6.1x greater than H_4_ reported previously. For LCL_AFR, 17 out of 34 hypotheses were not reported in OpenTargets, including *ITM2* and height^47^ (H_4_=0.97; see Suppl. Table 6). Of those reported previously, the average relative increase in P(H_4_) was 6.8x greater using LCL_AFR eQTLs in comparison to reported H_4_; with *EDC3* and height as example, we find H_4_ 2.3x greater in LCL_AFR compared to OpenTargets’ reporting of height^48^

Out of gene-phenotype pairs derived from eQTLs unique to African cohorts, only 2 were seen in both BRGR and LCL_AFR: *DRICH1* and eosinophil count in BCX (rs5759953 is eVariant in both cohorts) and *LLGL1* and height (rs112521610 eVariant in both). While colocalizations between *DRICH1* and lymphocyte, white blood cell and neutrophil count GWAS were observed n BCX^49^ using European-based eQTLs, colocalization with eosinophil count has not previously been reported. *LLGL1* eQTL signals have previously been observed to colocalize with GWAS of height, standing height and height at 10 years of age in Europeans^48–50^

## Discussion

The increased genetic diversity of individuals with African ancestry improves discovery power for regulatory mechanisms compared to Europeans. We performed eQTL studies in venous blood and LCLs in African and European cohorts and discovered that at the level of variants that passed QC, African ancestry cohorts have approximately 2x as many variants by tissue compared to European. We found enrichment of eQTLs in African compared to European cohorts: 3.7x more eQTLs in venous blood, and 5.6x more eQTLs in LCLs. While the discrepancy in number of variants or eQTLs in LCLs can be partially attributed to a nearly 2x greater sample size of the LCL_AFR compared to GEUVADIS, for venous blood PPMI sample size is slightly greater than BRGR.

Variant-for-variant, power increases with the increase in MAF from one population to another^51,52^. Here, 55.1% of eQTLs in BRGR and 61.7% in LCL_AFR are at least 3.16x more frequent as compared to PPMI or GEUVADIS. To facilitate comparisons between African and European eQTLs, we used a common human genome reference (GRCh38) and did not include the 297Mbp of genomic contigs identified in a large African WGS cohort^53^ that are absent from GRCh38. Using this updated reference may yield more ancestry-specific eQTLs than were discovered in the present study.

We avoided ancestry-based confounding in eQTL calling by using genetic and expression PCs, and showed that results were not affected by local ancestry. Interestingly, it was recently shown that DNA methylation QTLs are sensitive to local ancestry^54^, indicating that further work is needed to fully disentangle the contribution of local ancestry to different modalities of molecular phenotypes in *cis*

For variants with comparable allele frequencies in African and European cohorts, we quantified sharing of eQTLs using four measurements: exact (eGene, eVariant) pair, colocalization, replication and sharing at the level of eGenes. Within a tissue and across ancestry groups, we find QTLs shared under any definition to be low when considered as a fraction of total. Low eQTL sharing across tissues has previously been observed to hold generally in cross-tissue comparisons^55^, which we also observe in comparisons across tissues within an ancestry group.

Kachuri *et al* ^13^ stratified eQTLs into multiple tiers of sharing according to a decision tree, which does not readily allow direct comparison to other work. Nonetheless, of the genes they identified as heritable, 47.4% had no overlapping CS in their AFR_high_ cohort as compared to their AFR_low_ cohort. As only 9,609 genes met their definition of heritable, their conclusions broadly agree with ours. Recently, the AFGR^18^ cohort released a preprint studying the same HapMap samples comprising the LCL_AFR cohort, and report extensive sharing of eQTL effects with European LCL eQTLs. However, they measure sharing with mashr^56^, which specifically assesses whether a variant’s effect size is 0 or not in one or more groups. While interesting, this question is per-variant, and is only comparable to our exact match analysis, whereas our use of colocalization is targeted at identifying shared causal signals, and our replication analysis considers credible sets, both of which avoid confounding by unmodeled linkage disequilibrium. Despite the low eQTL sharing, shared eQTLs have similar effect sizes, agreeing with Stranger *et al.*^15^ that 34% of genetic effects are shared between ancestry groups with similar effect sizes. Beyond eQTLs, Kanai *et al* ^57^ studied the replication of fine-mapped GWAS variants (PIP > 0.9; “hits”) across three biobanks of different ancestry groups. Although their definitions of replication are both different from ours and multi-tiered, they find that 55% of hits meet their definition of fine-mapping-based replication, in broad agreement with our findings. Of the variants found not to replicate, they note that approximately 42% can be attributed to simply lower power in the other cohort(s), and another 42% can be attributed to differential allele frequencies. We speculate that the low replication rates observed in the present work may also be attributable to either lack of power, or differential allele frequency, especially given the pronounced differences in allele spectra we observe between ancestry groups (Fig. 2a, 2f).

Kachuri *et al* ^13^ found heritability of gene expression to differ by ancestry group, specifically their AFR_high_ group having more heritability on average as compared to their AFR_low_ group. We observed similar heritability differences when comparing BRGR and PPMI, and further showed that this comparison was confounded by differential genetic variance, and caution against overinterpretation of these findings in either the previous or current work.

In terms of gene mapping, we observed a meaningful increase in gene hypotheses when matching ancestries of GWAS and eQTL cohorts, and discovered colocalizations not previously reported. We also observed increased confidence in a number of previously reported gene hypotheses. While colocalization between GWAS and eQTL signals doesn’t necessarily imply that changes in gene expression levels of the eGene mediate genetic effects on disease in every instance^58^, the fact that only about 10% of index SNPs in the GWAS Catalog are located in the coding regions^55^ does mean that the use of eQTLs will remain a major strategy for nominating gene mapping hypotheses in a vast majority of GWAS loci. GWAS annotation analyses indicate the existence of novel gene-phenotype pairs discoverable only in African cohorts. We anticipate the release of the BRGR and LCL_AFR datasets will enable further research on these and other important questions pertaining to genetic regulation of gene expression in individuals of African descent.

## Online Methods

### Black Representation in Genomic Research (BRGR)

We recruited a cohort of 23andMe consented research participants who self-identified as being of African descent, were predicted to have ≥ 50% African ancestry by the 23andMe Ancestry Composition algorithm^26^, had no known blood related cancers or illness, and resided in the continental United States (see Suppl. Fig. 1 for details). WGS was performed on biobanked DNA from saliva samples of BRGR participants, and a venous blood sample was collected for RNA-seq. RNA extraction of 787 venous blood samples that were collected from 23andMe research participants using PAXgene blood RNA tubes was performed at the New York Genome Center (New York, NY). 2 samples were dropped due to contamination and an additional 14 were removed due to sample swaps. DNA extracted from blood cells in saliva samples of 23andMe customers was whole-genome sequenced at the Broad Institute (Cambridge, MA) with aligned CRAMs produced using their standard pipeline^59^. The average passing aligned read depth was 22.5x in the BRGR cohort. Randomization of BRGR WGS samples was performed to prevent batch effects using the blockTools R library^60^. 976 subjects had usable WGS samples. A total of 737 individuals from this cohort had saliva and venous blood samples of sufficiently high quality for downstream WGS and RNA-seq, respectively (see Suppl. Table 1 for details).

### Ethical approval

All biological sample collection was performed in accordance with the terms of informed consents and under an IRB approved protocol. Participants provided informed consent and participated in the research under a research protocol reviewed and approved by an external AAHRPP-accredited IRB, Ethical and Independent Review Services (www.eandireview.com). Participants consented to sharing of genetic and transcriptomic data via the NIH Database of Genotypes and Phenotypes (dbGaP).

### LCL_AFR

Lymphoblastoid cell lines (LCLs) from 660 individuals from the 1000 Genomes Project African superpopulation (all except HG02756, which is no longer available) were thawed and clones expanded at the Coriell Institute for Medical Research (Camden, NJ). This included individuals from the Yoruban, Esan, Gambian, Mende and Luhya continental African populations (including all YRI samples from the GEUVADIS project), as well as admixed African-American individuals from the U.S. Southwest and Afro-Caribbean individuals from Barbados (see Suppl. Table 3 for details). RNA from LCLs was extracted at the Coriell Institute. The final sample count was 659.

### RNA library preparation and sequencing

BRGR and LCL_AFR samples that met the following QC criteria: (1) minimum of 2µg total DNase-treated RNA, (2) absorbance values of OD260/280 ≥ 1.9 and (3) BioAnalyzer RIN value ≥ 8, were fragmented to 350bp average fragment length and prepared for sequencing using mRNA TruSeq Stranded kits (Illumina, San Diego, CA). Library preparation and RNA-sequencing of paired end 2×100bp reads was performed on Illumina NovaSeq sequencers at the New York Genome Center (New York, NY) to an average coverage of 60M reads.

### Parkinson Progression Marker Initiative (PPMI)

The PPMI^22,29^ cohort contains WGS and a series of functional measurements performed during the course of progression of Parkinson disease. We downloaded RNA-seq data and VCF files (Tier 2 Data) from the Image and Data Archive run by the Laboratory of Neuro Imaging (LONI) at the USC Mark and Mary Stevens Neuroimaging and Informatics Institute. For each sample, we took the RNA-seq from the earliest time point. Average coverage was about 100M reads per sample. The 1,379 initial samples as downloaded from USC were predominantly European (μ^European^=0.97±0.07), with the plurality of local European ancestry being Ashkenazi Jewish (μ^Ashkenazi^=0.36±0.48), followed by British and Irish (μ^British-Irish^=0.22±0.03). 1,238 samples were of broadly European ancestry.

Relatedness was highest in individuals of Ashkenazi descent (defined as μ^Ashkenaz^≥0.25) with 49.2% of pairwise comparisons having at least 0.05% of their genome that is IBD (Suppl. Fig. 2, Suppl. Table 7). In order to remove a potential bias in relatedness and heritability calculations due to including individuals from this founder population, and to standardize sample numbers in BRGR and PPMI cohorts so we remove an obvious confounder in eQTL statistical discovery power, we have excluded the 481 Ashkenazi Jewish individuals from our PPMI cohort. After removing 5 additional samples with incomplete gene expression data, the final sample count was 752.

### GEUVADIS

We downloaded RNA-seq data for the 358 European ancestry samples from the Geuvadis Consortium from https://www.ebi.ac.uk/biostudies/arrayexpress/studies/E-GEUV-1. Average coverage was about 20M reads per sample. LCL_AFR and GEUVADIS samples were part of the 1000 Genomes Project, and we downloaded high-coverage WGS data generated by the New York Genome Center^20^ from https://www.internationalgenome.org/data-portal/data-collection/30x-grch38.

### Variant calling

Variant calls were made using DeepVariant^61^ with joint calling performed by GLnexus^62,63^. The 1000 Genomes Project WGS samples (LCL_AFR and GEUVADIS cohorts) were processed with DeepVariant-0.8.0 and GLNexus-1.2.3 while for BRGR we used DeepVariant-1.1.0 and GLNexus-1.2.7. For PPMI, variant call files were downloaded through the LONI portal; variant calling in this cohort has been previously described^64^. Multi-allelic sites were split into individual alleles using `bcftools norm -m -any`. Genotypes with GQ<20 were set to missing, then variants were excluded where any of: >20% of genotypes missing, had no genotype with an alternative allele present or HWE exact test p-value<10^-50^

### Ancestry inference

To identify the ancestral origins of chromosomal segments across cohorts, we performed local ancestry inference using 23andMe’s Ancestry Composition^26^ Ancestry Composition uses support vector machine classifiers to assign one of 45 fine-scale ancestry populations to locally phased 300-SNP windows based on 541,948 SNPs. These preliminary assignments are next processed with an autoregressive pair hidden Markov model that smooths and corrects any phasing errors. The resulting posterior probabilities are recalibrated with an isotonic regression model. Local ancestry segments within each individual are finally summarized to produce a genome-wide proportion. For the purpose of this study, any population assignment with a mean less than 5% in either PPMI, African LCL, or BRGR was binned into a trace ancestry category. Finally, we performed independent randomization tests on 18 relevant ancestry populations to determine if the BRGR cohort ancestral representation is significantly different from that of a larger subset of individuals of African American participants. In each case, we performed a randomization test by randomly sampling (with replacement) 1000 23andMe research participants who identified as African-American based on survey answers and have >50% African ancestry (n=203,916) across 1000 iterations. For each iteration, we calculated the difference in mean genome-wide ancestry proportions between BRGR and the initial random subset (Δμ _Ancestry_^1^ ), then created two additional cohorts of 1000 individuals each by randomly sampling individuals from both starting cohorts and determined the difference in the means of these second cohorts ( Δμ _Ancestry_^2^ ). We determined p-values as the number of times that Δμ _Ancestry_^2^ was greater than or equal to Δμ _Ancestry_ out of the 1000 iterations.

### Identity-by-descent (IBD)

To determine relatedness between individuals we calculated the amount of DNA that is identical-by-descent between all pairs of individuals using phase-aware templated positional Burrows-Wheeler transform IBD detection (TPBWT-IBD)^65^. TPBWT-IBD was performed on all pairwise combinations of individuals across the PPMI, African LCL, and BRGR cohorts and a subset of additional 1000 Genomes Project^19^ and Human Genome Diversity Cell Line Panel (HGDP)^66^ populations. To maximize the accuracy of IBD detection, we used default parameters on an optimized set of 541,948 SNPs and retained IBD segments ≥ 5 centimorgans (cM). Finally, to visualize fine-scale ancestry, we arranged individuals in a graph based on the total amount of IBD they share using the ForceAtlas2^28^ layout. Force Atlas is an algorithm that situates individuals (or nodes) in a graph using a physical magnetic model. In this case, individuals with more IBD sharing will be attracted to one another and individuals with less IBD sharing are repelled. ForceAtlas2 runs until balance between repulsion and attraction is achieved, essentially illustrating fine-structure of individuals using the total IBD shared.

### RNA-seq mapping

RNA-seq reads were aligned to the human reference genome GRCh38 using STAR^67^ with 2 passes. Quality control for technical factors was done with FastQC^68^ and MultiQC^69^, and sample swaps were checked for using verifyBamID^70^. Per sample strand orientation was verified using the `infer_experiment.py` module in RSeQC^71^. Gene-level expression was quantified by HTSeq^72^ (`--stranded=reverse`) using GENCODE^73,74^ v28 gene models. RNA-seq reads were aligned to the whole transcriptome, but subsequently all analyses here and throughout the manuscript were limited to protein-coding genes.

### RNA-seq normalization

RNA-seq datasets were filtered for genes where ≥20% samples had a CPM (count per million) ≥0.1. Genes passing this threshold were further normalized first by scaling by library size using edgeR^75,76^, then converting to log_2_ scale. Finally, for all genes with more than 95% of samples exhibiting normalized expression value less than 4 standard deviations away from the expression mean, samples were right-truncated to the 95%-ile of normalized expression values; genes with 5% or more samples exhibiting normalized expression greater than 4 standard deviations from the mean were discarded from downstream analysis.

### eQTL discovery

eQTL discovery was done using the susieR package^30^ with default parameters: maximum 10 independent signals per gene, sum of credible set-level posterior inclusion probabilities (“coverage”) of 0.95 and minimum within-credible-set correlation (“purity”) of 0.5. The analysis was restricted to protein-coding genes and variants within a ±1Mbp window centered on the tested gene’s transcription start site (TSS). We tested SNVs and indels ≤ 500bp that had in-sample MAF≥1% and missingness < 5%. Covariates used across all datasets for eQTL calling included age, sex and 10 genetic PCs. Expression PCs were used to adjust for hidden covariates in RNA-seq data. The number of PCs was selected per dataset using the elbow method^77^, leading to 18 expression PCs for BRGR, 31 for LCL_AFR, 19 for PPMI and 23 for GEUVADIS. BRGR also included unique read fraction, RNA integrity number^78^ and % African ancestry as inferred by 23andMe Ancestry Classifier model^26^ GTEx also included sequencing platform and cohort (post-mortem, organ donor or surgical) as covariates.

### African-American GWAS

To assess the utility of the eQTL datasets for annotating GWAS hits, we downloaded African-American GWAS summary statistics from the VA Million Veteran Program (specifically blood lipids^23^, VTE^24^ and T2D^25^), and GWAS of blood traits from the Blood Cell Consortium^14^ Summary of all included GWAS are shown in Suppl. Table 5. As these GWAS had moderate sample sizes and numbers of genome-wide significant hits, we did not fully condition association signals in these studies.

In addition, we ran a GWAS of height in 23andMe’s African-American cohort, using our standard GWAS pipeline as described previously^79^ In short, we compute association test results for the genotyped and the imputed SNPs. For tests using imputed data, we use the imputed dosages rather than best-guess genotypes. As standard, we include covariates for age, gender, the first 6 PC of genetic ancestry to account for residual population structure, and indicators for genotype platforms to account for genotype batch effects. The association test p-value we report is computed using a likelihood ratio test. Association tests are performed by linear regression. Results for the chrX are computed similarly, with male genotypes coded as if they were homozygous diploid for the observed allele. Height GWAS signals were fully conditioned using the step-down conditional process: for each association genome wide, we re-ran the association test with the top variant from the preceding step in the model as an additional covariate at each iteration. The process is repeated up to 20 times or until no association is detected at p-value≤10^-5^. All conditionally independent variants identified were then introduced in a joint model. At each iteration one of the variants is left out to compute conditional leave each out (CLEO) statistics to be used in downstream analysis.

### Variant-to-gene mapping

African-American GWAS hits were linked to eGenes via colocalization analysis. Approximate Bayes factors^80^ were derived directly from MVP GWAS marginal summary statistics as the GWAS were likely underpowered to confidently detect secondary signals. For 23andMe African American height, each marginal association was further analyzed for conditionally independent signals using conditional leave each out (CLEO) analysis, and ABFs were computed using these conditionally-resolved statistics. ABFs representing independent signals were colocalized against SuSiE-derived eGenes using the coloc R package function coloc.susie_bf, with default parameters. Gene-trait pairs with a posterior probability of colocalization H_4_ ≥ {0.5, 0.8} are reported as colocalizing.

### Local ancestry-based eQTL calling

Local ancestry was incorporated into cis-eQTL calling using the Tractor^34^ model. We used Tractor to estimate European and African ancestry-specific effects and p-values by including alternate allele counts for each ancestry into the model. This was compared to a “standard” generalized linear regression model which measured the alternate allele effect, regardless of haplotype ancestry. In both models, we tested all variants within a 1Mb cis-window of each gene with MAF ≥ 5% and MAC ≥ 10 for both ancestry tracts. We adjusted for covariates including age, sex, 10 genetic PCs, 35 PEER factors, and sequencing factors in both models and for the Tractor model we also adjusted for the number of African ancestry haplotypes per locus.

### Heritability analysis

GCTA GREML^33^ was used to estimate cis-heritability for significant eGenes in BRGR and PPMI, separately, for autosomal variants with MAF ≥ 0.01. The phenotype used for GREML analysis was the normalized expression residualized on age, sex, 10 PCs of genetic ancestry, 35 PEER factors and sequencing covariates. We identified 9,777 eGenes that showed significant (p-value < 0.05) heritability in both cohorts. To avoid inflating estimates of *h*^2^ through relatedness, we filtered each cohort to the subset of individuals for which pairwise IBD < 0.025. To identify a high-confidence gene set for comparing heritability between BRGR and PPMI, we first derived fine-mapped^30,81^ credible sets for eQTLs in each cohort.

We focused our comparison on the 183 genes which had significant heritability estimates in both cohorts, and for which all 95% credible sets colocalized^29^ across the two cohorts with P(H_12_) > 0.5. For each gene *g* in each cohort, we approximately quantified the genetic variation (V_g_) that contributes to gene expression as the sum of genotype variance across the index variants in BRGR of *k* credible sets as

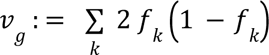

where *f*_k_ is the cohort-specific in-sample MAF for gene *g*. Here we assume that the genetic effect sizes are the same across cohorts and independent on variants so that, on average across the genes, we have 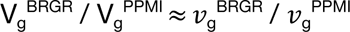. We use the BRGR index variants under the assumption that these colocalizing signals are shared causal eQTLs, and note that using PPMI index variants does not change our conclusions (Fig. 3). We compared the difference in heritability between BRGR and PPMI 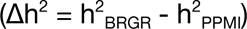 and the difference in v_g_ 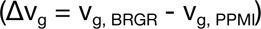.

For each cohort, we define the ratio of genetic to environmental variance explained for a given gene *g* as

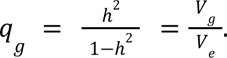

With terms 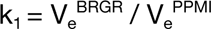 (ratio of environmental variances) and 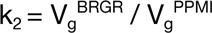 (ratio of genetic variances), for gene *g* we can relate BRGR and PPMI as

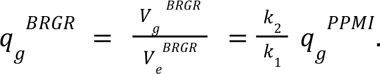

That is, the ratio of unexplained to explained variance for a gene in PPMI is proportional to the same quantity in BRGR. Under the assumption of approximately equal environmental variation, *k*_1_ ≈ 1, and

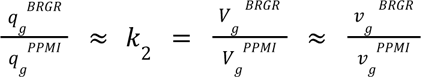

across genes broadly. This latter hypothesis can be tested by defining similar difference terms Δlog(q) and Δlog(v_g_), which are expected to be correlated. Relatedly, under this line of reasoning, log(q_BRGR_) = Δlog(v_g_) + log(q_PPMI_).

### OpenTargets Genetics access

OpenTargets Genetics v22.10 was accessed via API with queries against colocalization tables. We consider all gene-phenotype pairs reported in OpenTargets, regardless of eQTL or GWAS discovery cohorts, or tissues or cell types of discovery in the case of eQTLs. For a given eGene-phenotype pair discovered through one of the AFR cohorts, we search for similar OpenTargets Genetics phenotypes (i.e. MPV in BCX and ‘Mean platelet volume’ in UKB), and report the result from phenotype with the largest colocalization H_4_ with the eGene.

## Data availability

Consistent with the research consent provided, we made these datasets publicly available to all qualified researchers. RNA-seq and WGS data for Black Representation in Genomic Research is available as dbGaP study phs002969.v1.p1, and RNA-seq for the 1000 Genomes Project African superpopulation LCLs as SRA project PRJNA1108327. Upon publication the full summary statistics for the 23andMe African-American height GWAS and the BRGR eQTL study will be made available through 23andMe to qualified researchers under an agreement with 23andMe that protects the privacy of the 23andMe participants. Please visit https://research.23andme.com/collaborate/#dataset-access for more information and to apply to access the data. Upon publication genome-wide association summary statistics for the African LCL eQTL study will be freely available for download via https://www.internationalgenome.org.

## Acknowledgements

We thank the research participants and employees of 23andMe for making this work possible. We in particular thank the research participants of the Black Representation in Genomic Research (BRGR) study. BRGR participants provided informed consent and participated in the research under a protocol approved by the external AAHRPP-accredited IRB, Ethical & Independent Review Services (E&I Review).

The following members of the 23andMe Research Team contributed to this study: Stella Aslibekyan, Adam Auton, Elizabeth Babalola, Robert K. Bell, Jessica Bielenberg, Ninad S. Chaudhary, Zayn Cochinwala, Sayantan Das, Emily DelloRusso, Payam Dibaeinia, Sarah L. Elson, Nicholas Eriksson, Chris Eijsbouts, Teresa Filshtein, Pierre Fontanillas, Davide Foletti, Will Freyman, Zach Fuller, Julie M. Granka, Chris German, Éadaoin Harney, Alejandro Hernandez, Barry Hicks, David A. Hinds, M. Reza Jabal-Ameli, Ethan M. Jewett, Yunxuan Jiang, Sotiris Karagounis, Lucy Kaufmann, Matt Kmiecik, Katelyn Kukar, Alan Kwong, Keng-Han Lin, Yanyu Liang, Bianca A. Llamas, Aly Khan, Steven J. Micheletti, Matthew H. McIntyre, Meghan E. Moreno, Priyanka Nandakumar, Dominique T. Nguyen, Jared O’Connell, Steven J. Pitts, G. David Poznik, Alexandra Reynoso, Shubham Saini, Morgan Schumacher, Leah Selcer, Anjali J. Shastri, Jingchunzi Shi, Suyash Shringarpure, Keaton Stagaman, Teague Sterling, Qiaojuan Jane Su, Joyce Y. Tung, Susana A. Tat, Vinh Tran, Xin Wang, Wei Wang, Catherine H. Weldon, Amy L. Williams, Peter Wilton.

Some of the data used in the preparation of this manuscript was obtained in 2019 from the Parkinson’s Progression Markers Initiative (PPMI) database (www.ppmi-info.org/ access-data-specimens/download-data), RRID:SCR_006431. For up-to-date information on the study, visit www.ppmi-info.org. Our analysis used data from PPMI made available after either a pre-defined embargo or investigator submission of an associated cloud transfer request to the PPMI Data Access Committee. Protocol information for the Parkinson’s Progression Markers Initiative (PPMI) Clinical - Establishing a Deeply Phenotyped PD Cohort AM 3.2. can be found on protocols.io or by following this link: https://dx.doi.org/10.17504/ protocols.io.n92ldmw6ol5b/v2. PPMI – a public-private partnership – is funded by the Michael J. Fox Foundation for Parkinson’s Research, and funding partners; including 4D Pharma, Abbvie, AcureX, Allergan, Amathus Therapeutics, Aligning Science Across Parkinson’s, AskBio, Avid Radiopharmaceuticals, BIAL, BioArctic, Biogen, Biohaven, BioLegend, BlueRock Therapeutics, Bristol-Myers Squibb, Calico Labs, Capsida Biotherapeutics, Celgene, Cerevel Therapeutics, Coave Therapeutics, DaCapo Brainscience, Denali, Edmond J. Safra Foundation, Eli Lilly, Gain Therapeutics, GE HealthCare, Genentech, GSK, Golub Capital, Handl Therapeutics, Insitro, Jazz Pharmaceuticals, Johnson & Johnson Innovative Medicine, Lundbeck, Merck, Meso Scale Discovery, Mission Therapeutics, Neurocrine Biosciences, Neuron23, Neuropore, Pfizer, Piramal, Prevail Therapeutics, Roche, Sanofi, Servier, Sun Pharma Advanced Research Company, Takeda, Teva, UCB, Vanqua Bio, Verily, Voyager Therapeutics, the Weston Family Foundation and Yumanity Therapeutics.

## Author contributions

All authors contributed substantially to this manuscript.

## Supplementary Figures

**Supplementary Fig. 1.**
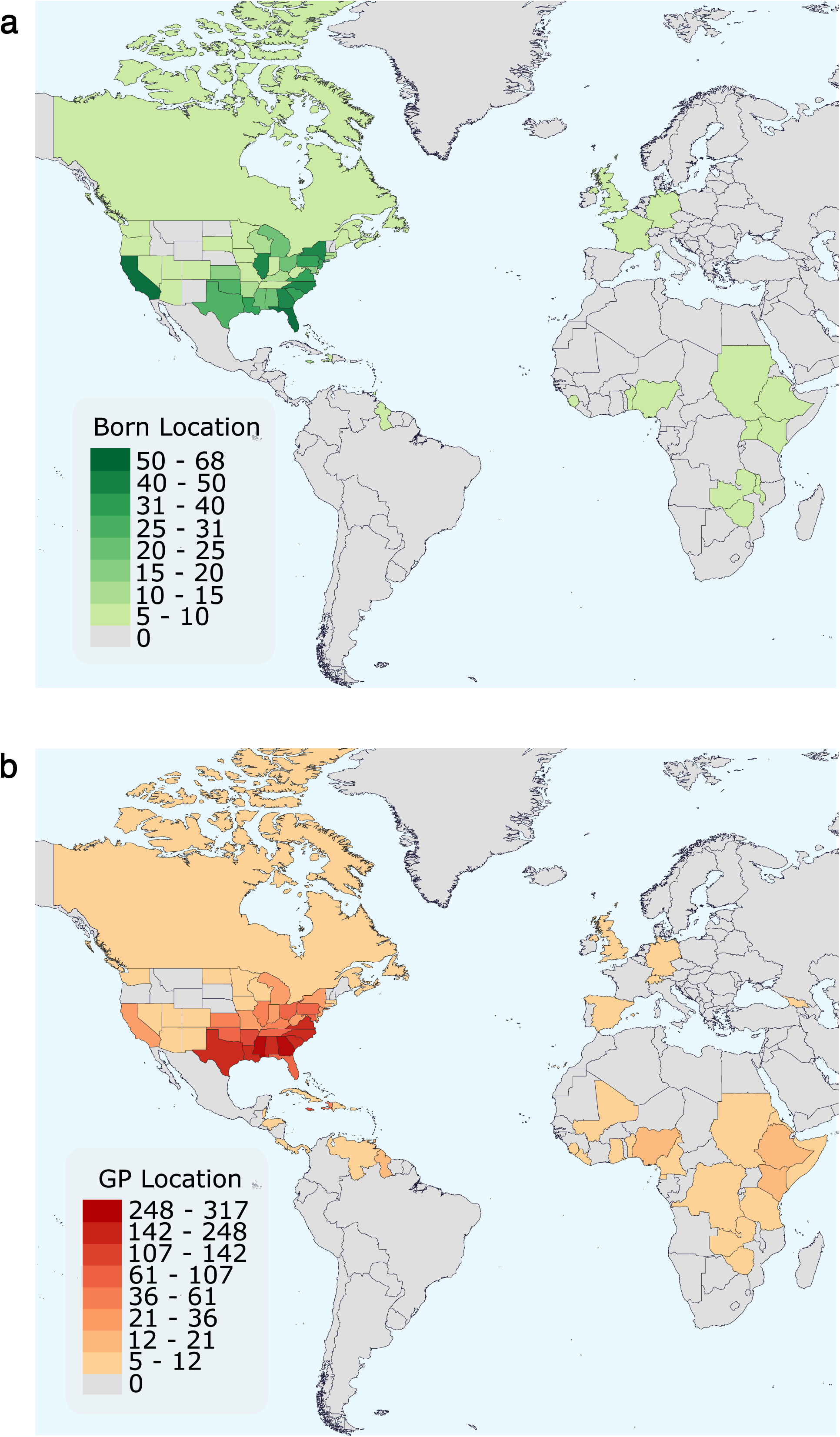
U.S. states / countries of birth for the Black Representation in Genomics Research cohort. Geographic representation of the BRGR cohort based on self-reported **(a)** birth locations (number of participants) and **(b)** birthplace of each grandparent (number of participants’ grandparents).

**Supplementary Fig. 2.**
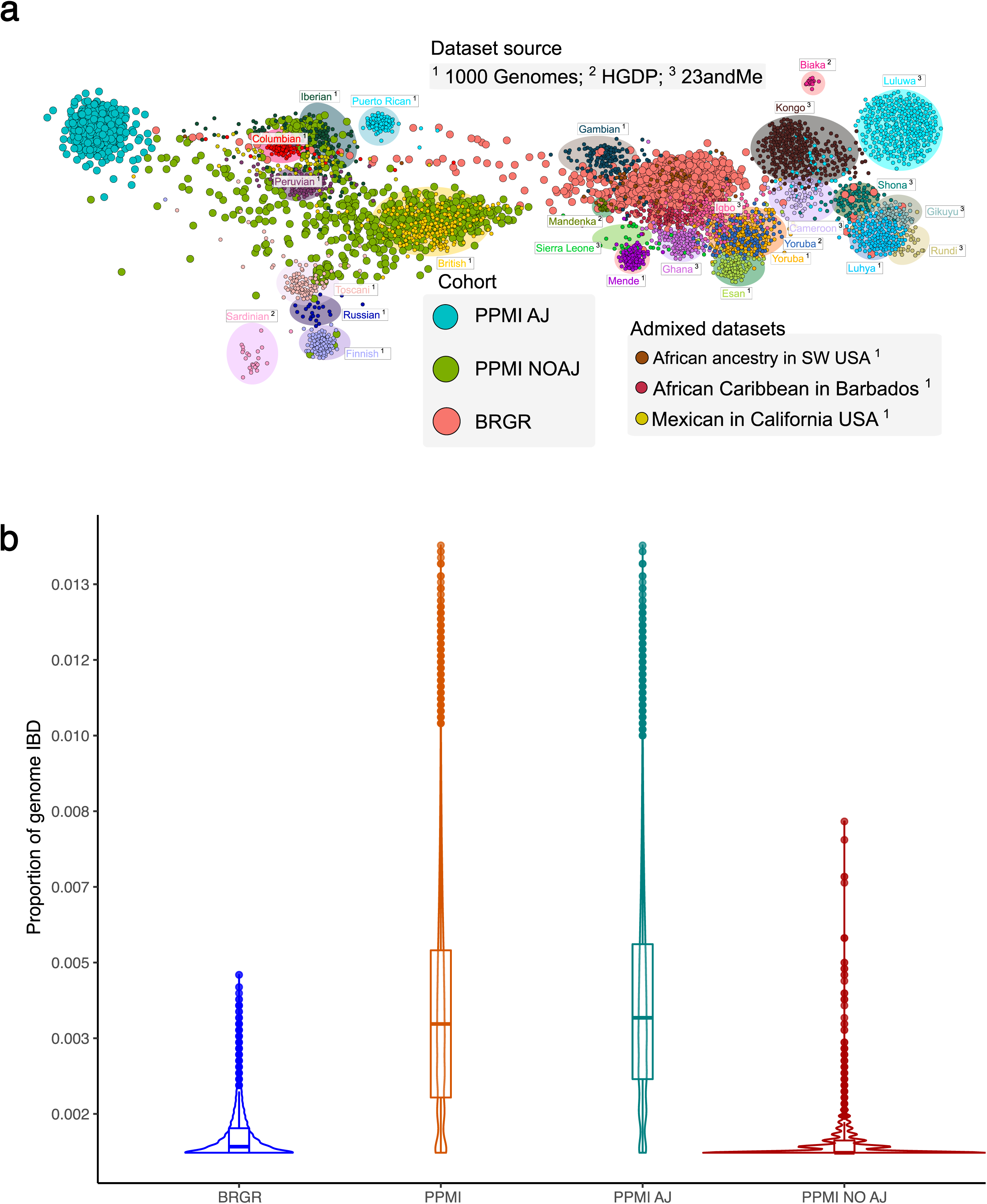
Summary of identical-by-descent (IBD) sharing in the BRGR and PPMI cohorts. **(a)** ForceAtlas2 network plot of individuals of African, European, and American descent using the total amount of DNA that is identical-by-descent between individuals. PPMI AJ individuals are those from the PPMI dataset that have ≥ 25% Ashkenazi Jewish ancestry, whereas PPMI NOAJ constitutes individuals with < 25 % Ashkenazi ancestry. **(b)** Difference in the proportion of genome that is identical by descent between BRGR, PPMI, and PPMI subset by individuals of Ashkenazi descent (PPMI AJ; ≥ 25% Ashkenazi ancestry) and individuals with < 25% Ashkenazi ancestry (PPMI NO AJ).

**Supplementary Fig. 3.**
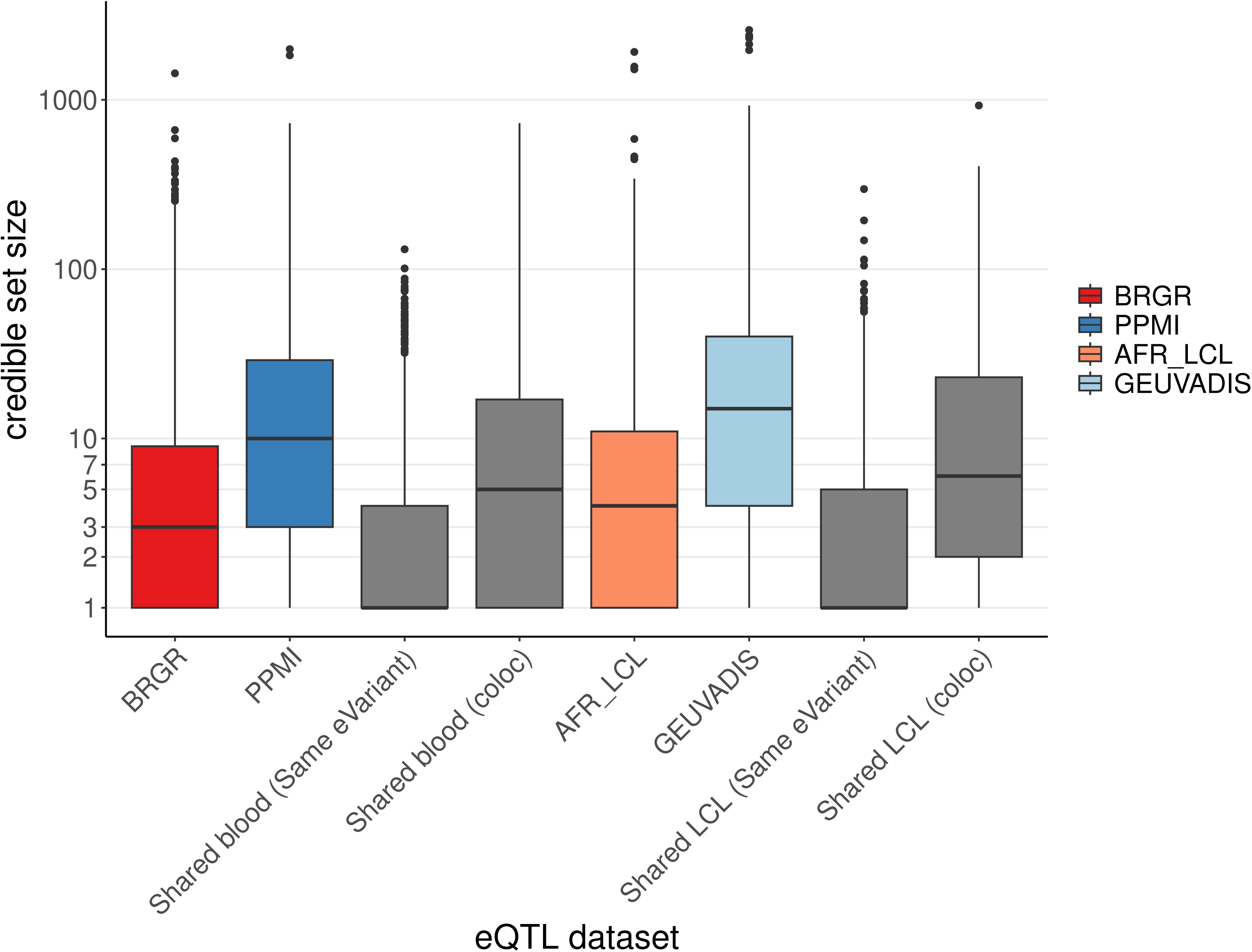
Distributions of credible set sizes in the four eQTL datasets and intersection of datasets.

**Supplementary Fig. 4.**
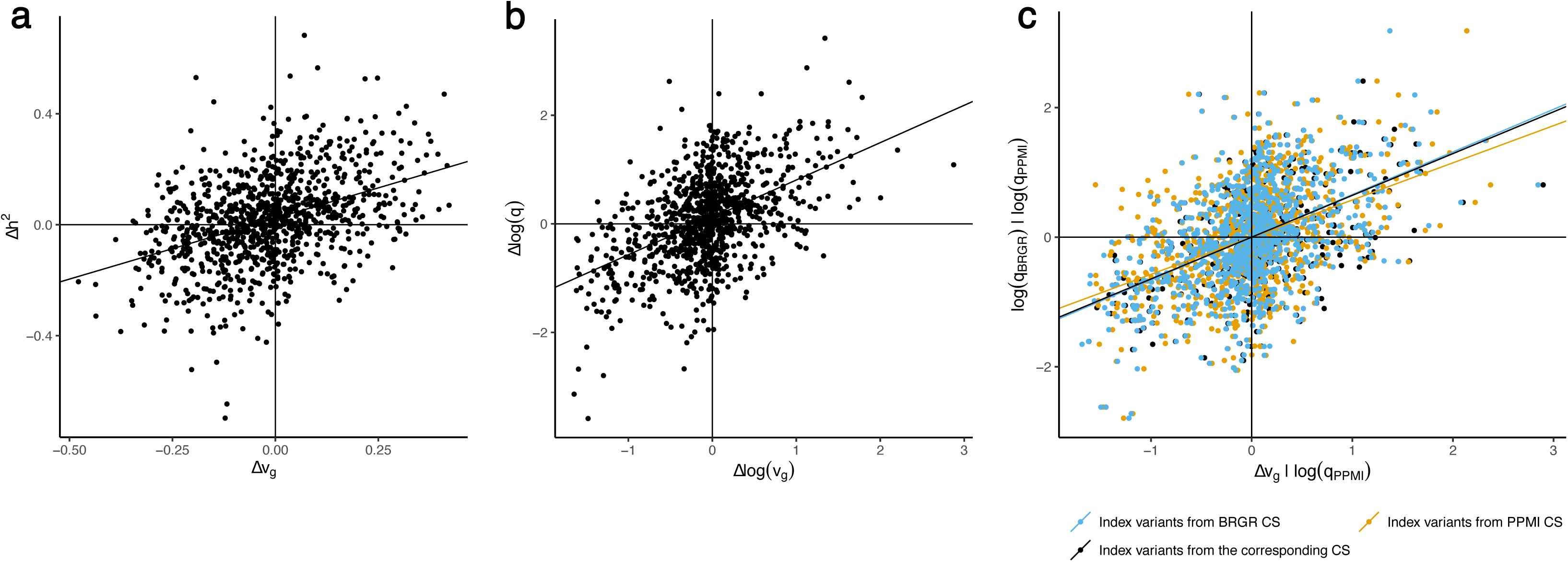
Explaining population differences in heritability using the differences in genetic diversity. Among the 183 genes whose credible sets are colocalized between BRGR and PPMI, we define the genetic diversity in three ways. Firstly, we assume that the causal eQTLs come from index variants of BRGR credible sets. Then the genetic diversity is calculated from the cohort-specific MAF of BRGR index variants (colored in blue). Similarly, we can define the genetic diversity based on the cohort-specific MAF of PPMI index variants (colored in yellow). Lastly, without determining the causal variants, we define the genetic diversity based on the index variants of the credible sets identified in the corresponding cohort (colored in black). **(a)** For each of the 183 genes, differences in genetic diversity Δv_g g, BRGR g, PPMI_ are shown on x-axis and differences in heritability 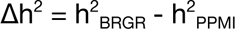 are shown on y-axis. The line is drawn from the linear fit Δh^2^∼ 1 + Δv_g_ **(b)** For each of the 183 genes, differences in logarithm of genetic diversity between BRGR and PPMI (Δlog(v_g_) = log(v_g, BRGR_) - log(v_g, PPMI_)) are shown on x-axis and differences in logarithm of transformed heritability (Δlog(q) = log(q_BRGR_) - log(q_PPMI_)) are shown on y-axis. The line is drawn from the linear fit Δlog(q) ∼ 1+ Δlog(v_g_). **(c)** For each of the 183 genes, the residuals of log(q_BRGR_) and Δlog(v_g_) after regressing out log(q_PPMI_) (intercept is also regressed out) are shown on y-axis and x-axis. The line is drawn from the linear fit residual(log(q_BRGR_)) ∼ residual(Δlog(v_g_)) whose slope is equal to the coefficient of Δlog(v_g_) in the linear fit log(q_BRGR_) ∼ 1 + log(q_PPMI_) + Δlog(v_g_).

**Supplementary Fig. 5.**
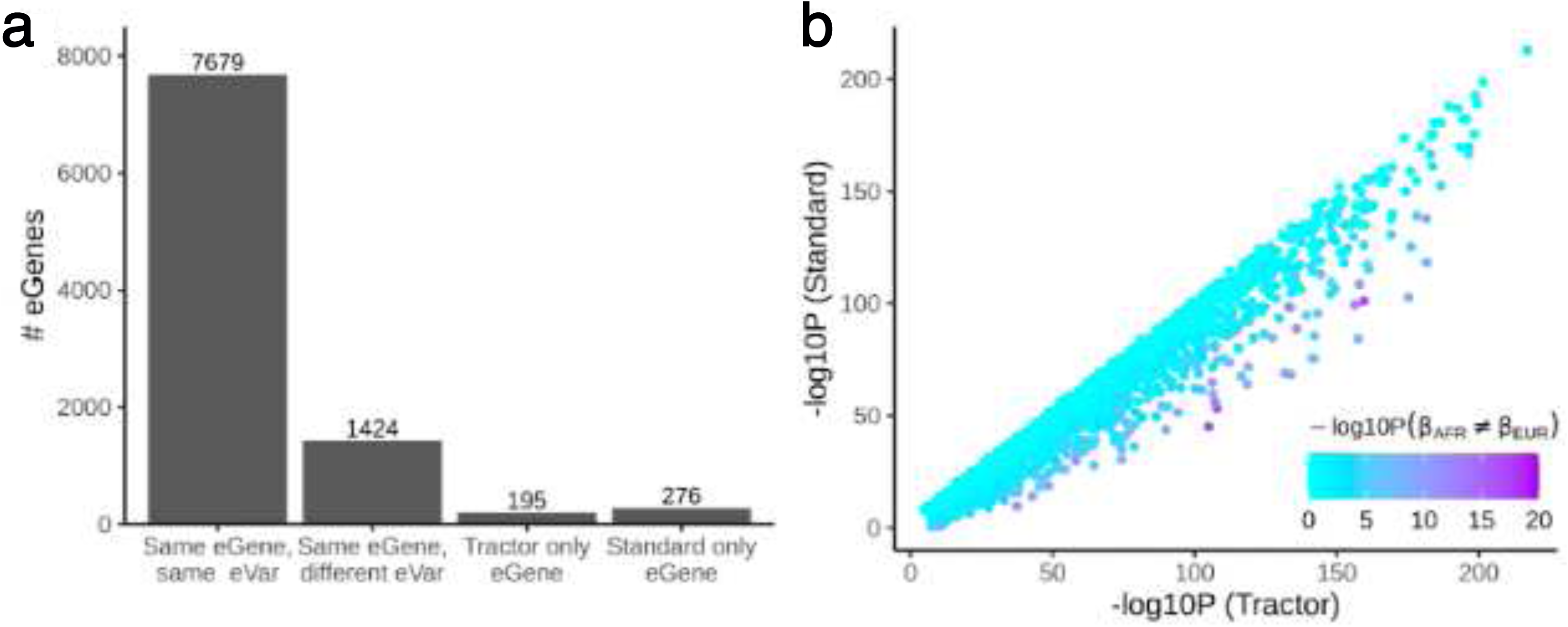
Local ancestry based cis-eQTL mapping using Tractor for 737 samples with admixed African ancestry from BRGR. Comparisons of genome-wide significant (p < 5×10^-8^) cis-eQTLs identified using a local-ancestry model, Tractor, or standard generalized linear model. **(a)** Shows the number of eGenes shared by or specific to each model. The majority of eGenes were identified in both models (“Same eGene, same eVar ’’ or “Same eGene, different eVar”). **(b)** For significant cis-eQTLs across both models, we compared p-values computed for each model. Ancestry-specific effects, estimated by Tractor, were also tested for significant differences. P-values were largely concordant between models for all cis-eQTLs; however, the p-value estimated by Tractor was slightly smaller for a subset of cis-eQTLs with heterogeneous effects between ancestries

## Supplementary Tables

**Supplementary Table 1.**
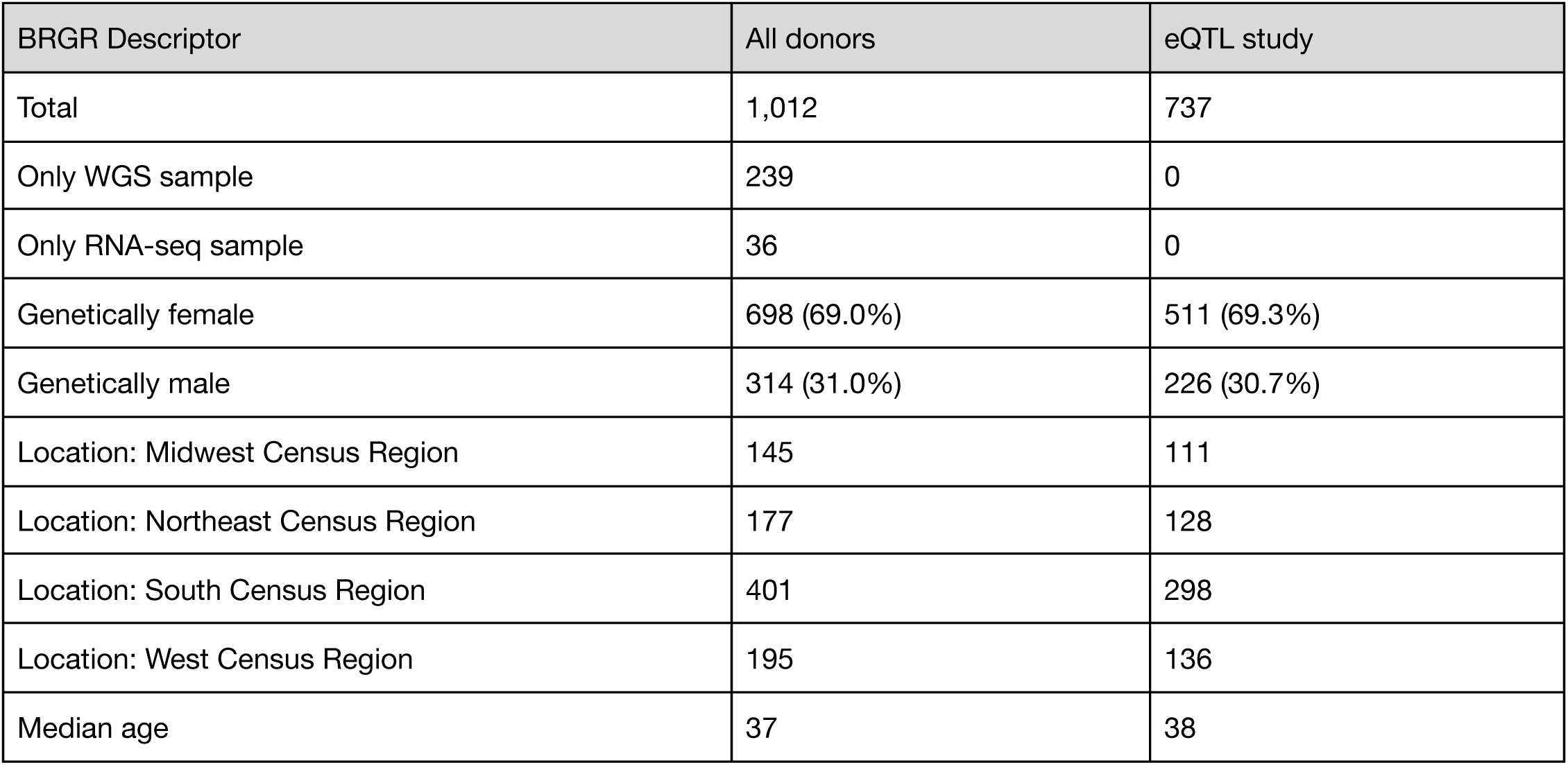
Summary of the Black Representation in Genomic Research (BRGR) cohort. The 737 donors that had both RNA-seq and WGS samples were included in the BRGR eQTL study. Location of sample collection was reported in terms of census regions as defined by the U.S. Census Bureau (https://www.census.gov/programs-surveys/economic-census/guidance-geographies/levels.html).

| BRGR Descriptor | All donors | eQTL study |
| --- | --- | --- |
| Total | 1,012 | 737 |
| Only WGS sample | 239 | 0 |
| Only RNA-seq sample | 36 | 0 |
| Genetically female | 698 (69.0%) | 511 (69.3%) |
| Genetically male | 314 (31.0%) | 226 (30.7%) |
| Location: Midwest Census Region | 145 | 111 |
| Location: Northeast Census Region | 177 | 128 |
| Location: South Census Region | 401 | 298 |
| Location: West Census Region | 195 | 136 |
| Median age | 37 | 38 |

**Supplementary Table 2.**
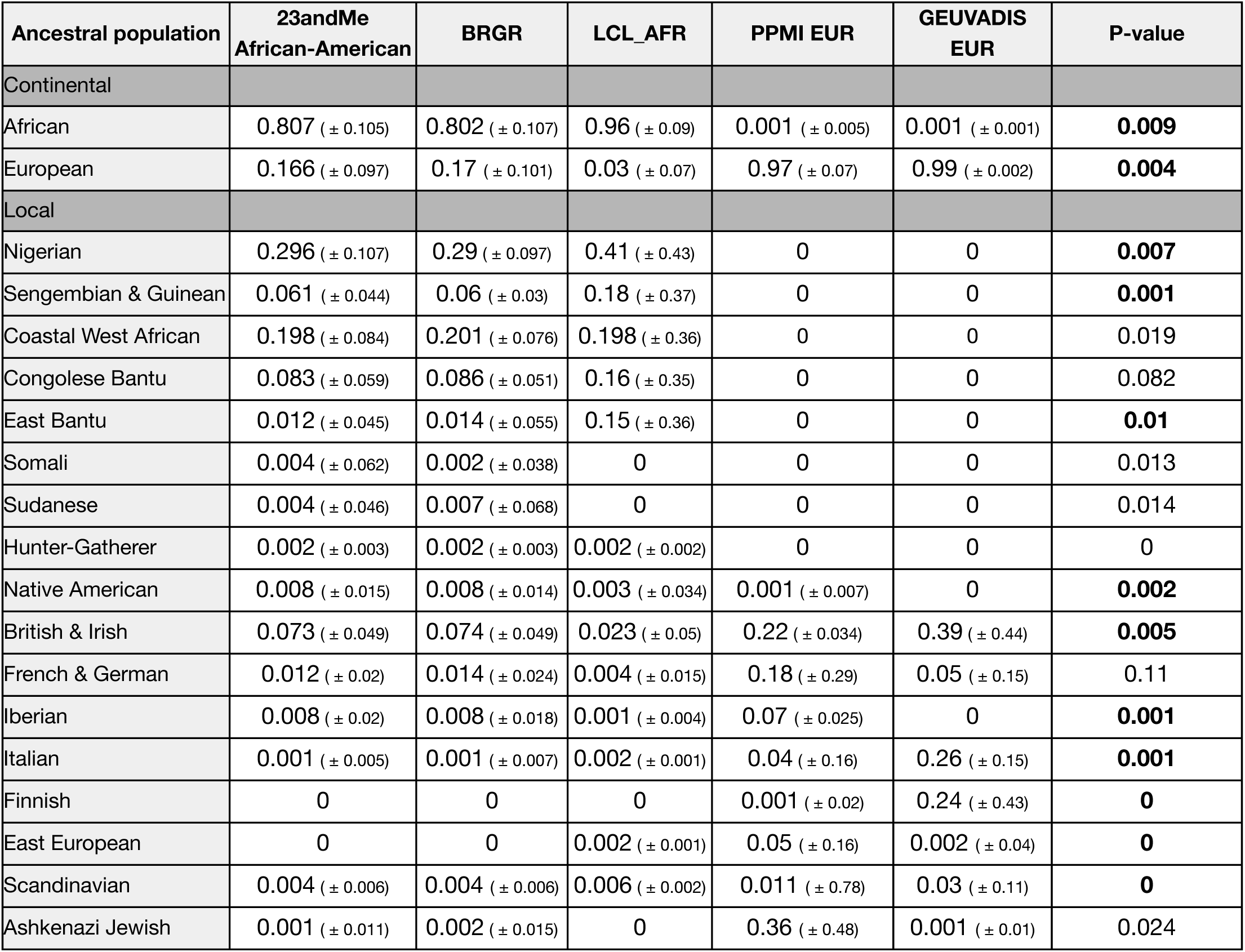
Mean genome-wide Ancestry Composition (± standard deviation) of represented broad and local populations in each cohort. *23andMe African-American* reference dataset consists of 203,937 research participants in the 23andMe database that self-identify as African-American and have ≥ 50% African ancestry. A Bonferroni corrected p-value < 0.012 indicates that the distribution of ancestry proportions between *23andMe African-American* and BRGR are indistinguishable based on a 1000-iteration randomization test.

| Ancestral population | 23andMe African-American | BRGR | LCL_AFR | PPMI EUR | GEUVADIS EUR | P-value |
| --- | --- | --- | --- | --- | --- | --- |
| Continental |  |  |  |  |  |  |
| African | 0.807 ($\pm 0.105$) | 0.802 ($\pm 0.107$) | 0.96 ($\pm 0.09$) | 0.001 ($\pm 0.005$) | 0.001 ($\pm 0.001$) | **0.009** |
| European | 0.166 ($\pm 0.097$) | 0.17 ($\pm 0.101$) | 0.03 ($\pm 0.07$) | 0.97 ($\pm 0.07$) | 0.99 ($\pm 0.002$) | **0.004** |
| Local |  |  |  |  |  |  |
| Nigerian | 0.296 ($\pm 0.107$) | 0.29 ($\pm 0.097$) | 0.41 ($\pm 0.43$) | 0 | 0 | **0.007** |
| Sengembian & Guinean | 0.061 ($\pm 0.044$) | 0.06 ($\pm 0.03$) | 0.18 ($\pm 0.37$) | 0 | 0 | **0.001** |
| Coastal West African | 0.198 ($\pm 0.084$) | 0.201 ($\pm 0.076$) | 0.198 ($\pm 0.36$) | 0 | 0 | 0.019 |
| Congolese Bantu | 0.083 ($\pm 0.059$) | 0.086 ($\pm 0.051$) | 0.16 ($\pm 0.35$) | 0 | 0 | 0.082 |
| East Bantu | 0.012 ($\pm 0.045$) | 0.014 ($\pm 0.055$) | 0.15 ($\pm 0.36$) | 0 | 0 | **0.01** |
| Somali | 0.004 ($\pm 0.062$) | 0.002 ($\pm 0.038$) | 0 | 0 | 0 | 0.013 |
| Sudanese | 0.004 ($\pm 0.046$) | 0.007 ($\pm 0.068$) | 0 | 0 | 0 | 0.014 |
| Hunter-Gatherer | 0.002 ($\pm 0.003$) | 0.002 ($\pm 0.003$) | 0.002 ($\pm 0.002$) | 0 | 0 | 0 |
| Native American | 0.008 ($\pm 0.015$) | 0.008 ($\pm 0.014$) | 0.003 ($\pm 0.034$) | 0.001 ($\pm 0.007$) | 0 | **0.002** |
| British & Irish | 0.073 ($\pm 0.049$) | 0.074 ($\pm 0.049$) | 0.023 ($\pm 0.05$) | 0.22 ($\pm 0.034$) | 0.39 ($\pm 0.44$) | **0.005** |
| French & German | 0.012 ($\pm 0.02$) | 0.014 ($\pm 0.024$) | 0.004 ($\pm 0.015$) | 0.18 ($\pm 0.29$) | 0.05 ($\pm 0.15$) | 0.11 |
| Iberian | 0.008 ($\pm 0.02$) | 0.008 ($\pm 0.018$) | 0.001 ($\pm 0.004$) | 0.07 ($\pm 0.025$) | 0 | **0.001** |
| Italian | 0.001 ($\pm 0.005$) | 0.001 ($\pm 0.007$) | 0.002 ($\pm 0.001$) | 0.04 ($\pm 0.16$) | 0.26 ($\pm 0.15$) | **0.001** |
| Finnish | 0 | 0 | 0 | 0.001 ($\pm 0.02$) | 0.24 ($\pm 0.43$) | **0** |
| East European | 0 | 0 | 0.002 ($\pm 0.001$) | 0.05 ($\pm 0.16$) | 0.002 ($\pm 0.04$) | **0** |
| Scandinavian | 0.004 ($\pm 0.006$) | 0.004 ($\pm 0.006$) | 0.006 ($\pm 0.002$) | 0.011 ($\pm 0.78$) | 0.03 ($\pm 0.11$) | **0** |
| Ashkenazi Jewish | 0.001 ($\pm 0.011$) | 0.002 ($\pm 0.015$) | 0 | 0.36 ($\pm 0.48$) | 0.001 ($\pm 0.01$) | 0.024 |

**Supplementary Table 3.** Summary of the 1000 Genomes Project samples included in the African ancestry superpopulation LCL eQTL cohort, and in the European component of GEUVADIS.

| Superpopulation | Code | Population | Females | Males | Both sexes |
| --- | --- | --- | --- | --- | --- |
| AFR | ACB | African Caribbeans in Barbados | 49 | 47 | 96 |
| AFR | ASW | Americans of African Ancestry in SW USA | 35 | 26 | 61 |
| AFR | ESN | Esan in Nigeria | 46 | 53 | 99 |
| AFR | GWD | Gambian in Western Divisions in the Gambia | 58 | 53 | 111 |
| AFR | LWK | Luhya in Webuye, Kenya | 55 | 44 | 99 |
| AFR | MSL | Mende in Sierra Leone | 43 | 42 | 85 |
| AFR | YRI | Yoruba in Ibadan, Nigeria | 56 | 52 | 108 |
| **AFR** | **Total** |  | **342 (51.8%)** | **318 (48.2%)** | **659** |
| EUR | CEU | CEPH | 44 | 45 | 89 |
| EUR | FIN | Finns | 56 | 36 | 92 |
| EUR | GBR | British | 43 | 43 | 86 |
| EUR | TSI | Toscani | 44 | 47 | 91 |
| **EUR** | **Total** |  | **187 (52.2%)** | **171 (47.8%)** | **358** |

**Supplementary Table 4.**
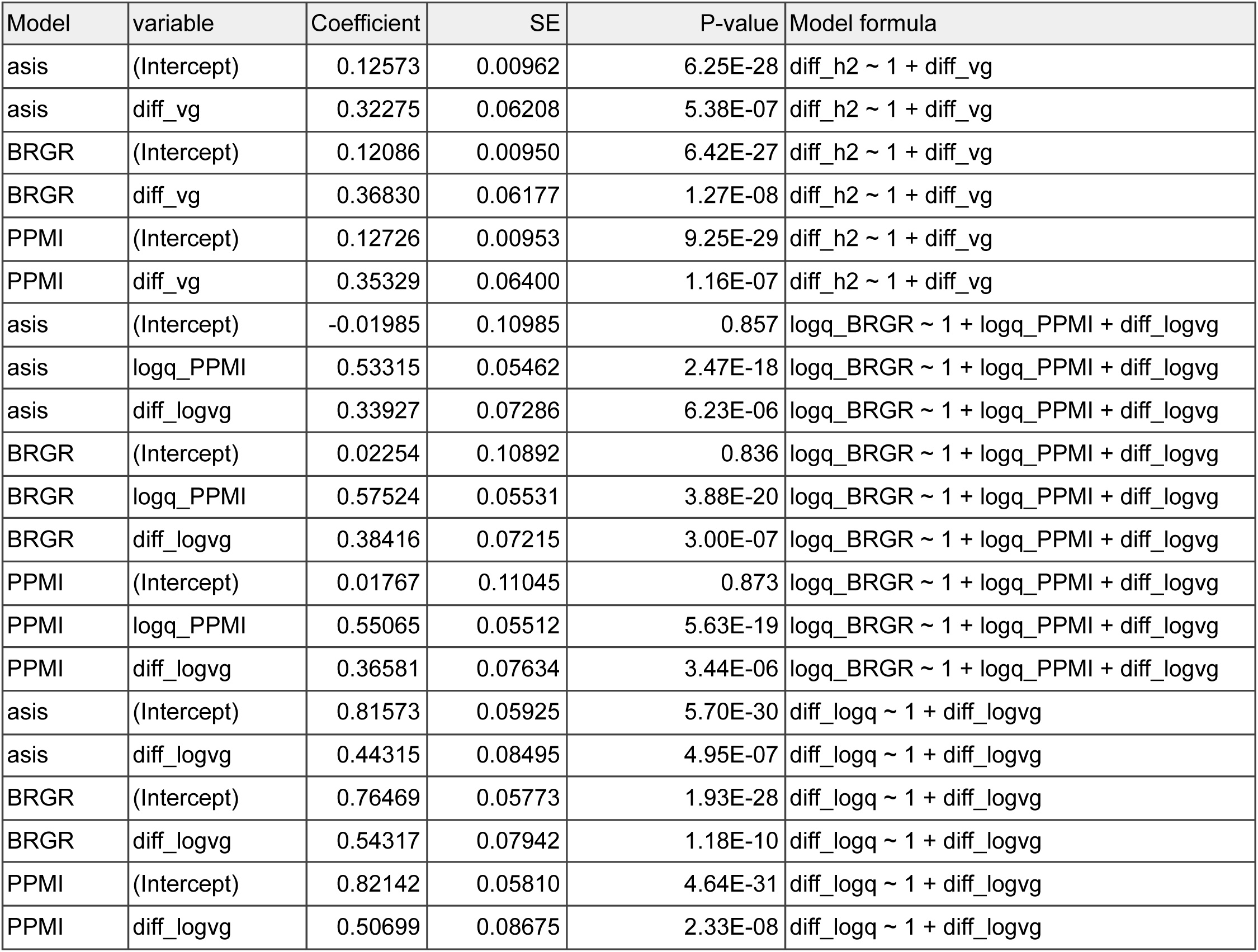
Coefficients of the regression analysis in testing the relation between differences in heritability and differences in genetic diversity. Assuming no differential causal effect and environmental noise, we derived a relation describing how genetic diversity differences result in differences in heritability: v_g, BRGR_ / v_g, PPMI_ ≈ q_BRGR_ / q_PPM_ or equivalently Δlog(v_g_) - Δlog(q) ≈ 0. This relation is examined in various regression analysis using the 183 genes whose credible sets are colocalized between BRGR and PPMI. The following linear regression analysis were performed: 1) Δh^2^ ∼ 1 + Δv_g_; 2) Δlog(q) ∼ 1 + Δlog(v_g_); 3) log(q_BRGR_) ∼ 1 + log(q_PPMI_) + Δlog(v_g_). The coefficients, standard errors, and p-values of these regressions are shown.

| Model | variable | Coefficient | SE | P-value | Model formula |
| --- | --- | --- | --- | --- | --- |
| asis | (Intercept) | 0.12573 | 0.00962 | 6.25E-28 | diff_h2 ~ 1 + diff_vg |
| asis | diff_vg | 0.32275 | 0.06208 | 5.38E-07 | diff_h2 ~ 1 + diff_vg |
| BRGR | (Intercept) | 0.12086 | 0.00950 | 6.42E-27 | diff_h2 ~ 1 + diff_vg |
| BRGR | diff_vg | 0.36830 | 0.06177 | 1.27E-08 | diff_h2 ~ 1 + diff_vg |
| PPMI | (Intercept) | 0.12726 | 0.00953 | 9.25E-29 | diff_h2 ~ 1 + diff_vg |
| PPMI | diff_vg | 0.35329 | 0.06400 | 1.16E-07 | diff_h2 ~ 1 + diff_vg |
| asis | (Intercept) | -0.01985 | 0.10985 | 0.857 | logq_BRGR ~ 1 + logq_PPMI + diff_logvg |
| asis | logq_PPMI | 0.53315 | 0.05462 | 2.47E-18 | logq_BRGR ~ 1 + logq_PPMI + diff_logvg |
| asis | diff_logvg | 0.33927 | 0.07286 | 6.23E-06 | logq_BRGR ~ 1 + logq_PPMI + diff_logvg |
| BRGR | (Intercept) | 0.02254 | 0.10892 | 0.836 | logq_BRGR ~ 1 + logq_PPMI + diff_logvg |
| BRGR | logq_PPMI | 0.57524 | 0.05531 | 3.88E-20 | logq_BRGR ~ 1 + logq_PPMI + diff_logvg |
| BRGR | diff_logvg | 0.38416 | 0.07215 | 3.00E-07 | logq_BRGR ~ 1 + logq_PPMI + diff_logvg |
| PPMI | (Intercept) | 0.01767 | 0.11045 | 0.873 | logq_BRGR ~ 1 + logq_PPMI + diff_logvg |
| PPMI | logq_PPMI | 0.55065 | 0.05512 | 5.63E-19 | logq_BRGR ~ 1 + logq_PPMI + diff_logvg |
| PPMI | diff_logvg | 0.36581 | 0.07634 | 3.44E-06 | logq_BRGR ~ 1 + logq_PPMI + diff_logvg |
| asis | (Intercept) | 0.81573 | 0.05925 | 5.70E-30 | diff_logq ~ 1 + diff_logvg |
| asis | diff_logvg | 0.44315 | 0.08495 | 4.95E-07 | diff_logq ~ 1 + diff_logvg |
| BRGR | (Intercept) | 0.76469 | 0.05773 | 1.93E-28 | diff_logq ~ 1 + diff_logvg |
| BRGR | diff_logvg | 0.54317 | 0.07942 | 1.18E-10 | diff_logq ~ 1 + diff_logvg |
| PPMI | (Intercept) | 0.82142 | 0.05810 | 4.64E-31 | diff_logq ~ 1 + diff_logvg |
| PPMI | diff_logvg | 0.50699 | 0.08675 | 2.33E-08 | diff_logq ~ 1 + diff_logvg |

**Supplementary Table 5.** Summary of African-American GWAS used and detailed summary of variant-to-gene mapping. MVP - the VA Million Veteran Program; BCX - Blood Cell Consortium.

| Source | Trait | dbGaP analysis accession | Cases | Controls | GWAS hits | N coloc ($H_4 \geq .8$) | | | | N coloc ($H_4 \geq .5$) | | | |
| --- | --- | --- | --- | --- | --- | --- | --- | --- | --- | --- | --- | --- | --- |
|  |  |  |  |  |  | BRGR | PPMI | LCL_AFR | GEUVADIS | BRGR | PPMI | LCL_AFR | GEUVADIS |
| MVP | blood lipids - HDL | pha004827.1 | 57,332 |  | 63 | 2 | 1 | 3 | - | 4 | 1 | 3 | - |
| MVP | blood lipids - LDL | pha004830.1 |  |  | 53 | 1 | - | - | - | 1 | - | - | - |
| MVP | blood lipids - total cholesterol (TC) | pha004833.1 |  |  | 77 | 1 | - | - | - | 2 | - | - | - |
| MVP | blood lipids - triglycerides (TG) | pha004836.1 |  |  | 47 | - | 1 | 1 | - | - | 1 | 1 | 1 |
| MVP | venous thromboembolism (VTE) | pha004962.1 | 2,261 | 49,400 | 16 | 1 | - | 1 | - | 1 | - | - | - |
| MVP | type 2 diabetes (T2D) | pha004943.1 | 24,646 | 31,446 | 56 | 1 | - | - | - | 1 | - | - | - |
| **MVP** | **total across all traits** |  |  |  | **312** | **6** | **2** | **4** | **0** | **9** | **2** | **4** | **1** |
| BCX | red blood cell count (RBC count) |  | Up to 15,171.<br>Varies variant by variant depending on how many studies were included in a meta-analysis. |  | 32 | 2 | - | - | - | 2 | 1 | 1 | - |
| BCX | hemoglobin concentration (HGB) |  |  |  | 22 | - | - | 1 | - | - | - | 1 | - |
| BCX | hematocrit (HCT) |  |  |  | 17 | - | - | - | - | - | - | - | - |
| BCX | mean corpuscular hemoglobin (MCH) |  |  |  | 19 | - | - | - | 1 | - | - | - | 1 |
| BCX | mean corpuscular volume (MCV) |  |  |  | 21 | 2 | 1 | 1 | 1 | 2 | 2 | 1 | 1 |
| BCX | mean corpuscular hemoglobin concentration (MCHC) |  |  |  | 39 | 3 | - | 1 | - | 7 | 1 | 4 | - |
| BCX | RBC distribution width (RDW) |  |  |  | 27 | 2 | - | - | - | 2 | - | - | - |
| BCX | total white blood cell count (WBC count) |  |  |  | 46 | 5 | - | - | - | 6 | - | - | - |
| BCX | neutrophil count (Neutro) |  |  |  | 37 | 4 | 1 | - | - | 4 | 1 | - | - |
| BCX | lymphocyte count (Lympho) |  |  |  | 29 | 1 | - | 2 | - | 1 | - | 2 | - |
| BCX | monocyte count (Mono) |  |  |  | 33 | 3 | 2 | - | - | 3 | 2 | 1 | - |
| BCX | basophil count (Baso) |  |  |  | 35 | - | - | - | - | - | - | - | - |
| BCX | eosinophil count (Eosin) |  |  |  | 28 | 2 | 2 | 2 | 3 | 2 | 3 | 2 | 3 |
| BCX | platelet count (PLT count) |  |  |  | 38 | 4 | 2 | 3 | 2 | 7 | 3 | 4 | 2 |
| BCX | mean platelet volume (MPV) |  |  |  | 38 | 4 | 3 | 3 | 1 | 5 | 3 | 3 | 2 |
| **BCX** | **total across all traits** |  |  |  | **461** | **32** | **11** | **13** | **8** | **41** | **16** | **19** | **9** |
| **23andMe** | **height** | **NA** | **273,215** |  | **803** | **37** | **28** | **36** | **13** | **69** | **53** | **64** | **24** |

**Supplementary Table 6 | Variant-to-gene hypothesis derived from eQTLs unique to African ancestry cohorts.** MVP - the VA Million Veteran Program; BCX - Blood Cell Consortium.

**Supplementary Table 7.** Number of relationships based on the proportion of the genome that is identical-by-descent (IBD) between all pairwise combinations of individuals in each cohort.

|  | N pairwise relationships |  |  |  |
| --- | --- | --- | --- | --- |
| IBD Range<br>(proportion genome) | PPMI | PPMI (with AJ) | BRGR | Typical relationship(s) in range |
| < 0.001 | 575,626 | 118,123 | 1,015,119 | Unrelated |
| 0.001-0.005 | 4,841 | 84,903 | 8,959 | 4th, 5th and distant cousins |
| 0.005-0.01 | 92 | 28,924 | 27 | 3rd cousins |
| 0.01-0.025 | 17 | 770 | 11 | Half 2nd cousins |
| 0.025-0.05 | 13 | 5 | 5 | 2nd cousins |
| 0.05-0.1 | 5 | 5 | 5 | Great grandparents |
| 0.1-0.2 | 12 | 5 | 5 | 1st cousins |
| 0.2-0.3 | 6 | 12 | 5 | Grandparents, avuncular |
| 0.3-1 | 32 | 59 | 8 | Full siblings, parents, twins |

